# MES: A Multi-Agent Evidence Synthesis System for Medical Decision-Making

**DOI:** 10.64898/2026.09.15.26362921

**Authors:** Haoyang Li, Weishen Pan, Suraj Rajendran, Chengxi Zang, Judy Gabeskiria, Valencia Liu, Vivian Feng, Mahitha Penmetsa, Lydia Lin, Atindra Bhalchandra Jayakar, Ayham Boucher, Edward J. Schenck, He S. Yang, Fei Wang

**Affiliations:** Department of Population Health Sciences, Weill Cornell Medicine, New York, NY, USA; Tri-Institutional Computational Biology & Medicine Program, Cornell University, New York, NY, USA; Weill Cornell Medicine, New York, NY, USA; Cornell AI Innovation Hub, Cornell University, Ithaca, NY, USA; Division of Pulmonary and Critical Care Medicine, Department of Medicine, New York-Presbyterian Hospital-Weill Cornell Medical Center, Weill Cornell Medicine, New York, NY, USA; Department of Pathology and Laboratory Medicine, Weill Cornell Medicine, New York, NY, USA

## Abstract

Medical evidence synthesis increasingly requires published studies, real-world clinical data and structured biomedical knowledge, yet most automated systems remain centered on literature retrieval and summarization. Here we present MES, a multi-agent framework for source-grounded medical evidence synthesis. MES coordinates six specialized agents to decompose clinical questions, retrieve literature and trial evidence, generate question-specific RWE by constructing and analyzing real-world cohorts, query biomedical knowledge graphs, integrate quantitative findings and screen generated claims against their cited evidence. The framework uses evidence-based medicine taxonomies to clarify underspecified questions, adapts evidence use when sources are absent or discordant and reports unresolved gaps rather than forcing consensus. MES also provides claim-level provenance tracking and cross-agent consistency checking, allowing final reports to distinguish trial evidence, real-world associations and knowledge-graph support. We evaluated MES in two clinical use cases and across 144 clinical queries spanning six evidence-based medicine categories. MES generated structured reports that preserved source traceability, identified cross-source disagreement and communicated uncertainty across diverse clinical question types. MES provides an auditable framework for organizing, testing and contextualizing heterogeneous clinical evidence while keeping the evidentiary basis of each conclusion explicit.

## Introduction

Evidence-based medicine (EBM)^1,2^ depends on connecting clinical questions with the best available evidence and judging how far that evidence applies to a patient, population or decision. Relevant evidence is no longer limited to randomized controlled trials (RCTs)^3^ and systematic reviews. Real-world evidence (RWE)^4^ can be generated from real-world data sources including electronic health records (EHRs)^5,6^, administrative claims, disease and product registries, patient-reported surveys, wearable sensors and home monitoring devices.^7–9^ Published observational studies provide additional evidence about care outside randomized settings, while structured biomedical knowledge graphs organize relationships among diseases, drugs, genes, pathways and phenotypes. These evidence sources answer different parts of a clinical question. RCTs estimate effects under defined study conditions, whereas real-world analyses can characterize outcomes in routine care and in populations that may be under-represented in trials. Knowledge graphs provide complementary mechanistic and ontological context. The central challenge for evidence synthesis is therefore not only to find evidence, but also to distinguish evidence types, account for differences in population and study design and make the provenance and applicability of each conclusion explicit.

Systematic reviews and meta-analyses^10^ remain reference standards for many evidence-synthesis tasks, but their fundamental scope is the synthesis of evidence that has already been generated and made available. For many clinical questions, especially those involving narrowly defined subgroups, safety signals, local practice patterns, contemporary treatment pathways or prognosis under routine care, published evidence may be sparse or only indirectly applicable. In such settings, identifying the literature gap is informative but does not itself characterize outcomes in the population or setting of interest. Analysis of underlying real-world clinical data can provide complementary, question-specific empirical evidence, including evidence reflecting more local or recent practice. Such analyses, however, introduce their own limitations, including confounding, missingness, measurement error and uncertain generalizability. Clinical evidence synthesis must therefore distinguish evidence generated under different designs, expose these limitations and preserve a traceable link between each conclusion and its supporting source.

Large language models (LLMs)^11,12^ have accelerated biomedical search, screening, extraction and summarization.^13–15^ Recent systems and tools^16–19^, including OpenEvidence, can retrieve cited biomedical literature, synthesize findings across published studies and answer clinical questions using both randomized and observational evidence. A distinct capability beyond literature retrieval, however, is to generate question-specific RWE directly from underlying patient-level clinical data when the available literature does not adequately address the target population, setting or decision. This requires translating a clinical question into computable cohort definitions, selecting an appropriate analytical design and executing the analysis rather than retrieving an already published RWE study. More broadly, three challenges remain for automated clinical evidence synthesis. First, newly generated RWE must remain distinguishable from retrieved published evidence, including randomized trials and observational studies, and must be interpreted in light of its specific risks of bias, confounding and limited transportability. Second, clinical questions may require decomposition, clarification and iterative follow-up rather than treatment as fixed prompts. Third, citation does not by itself establish factual support: a generated statement may cite a relevant source while overstating its result, omitting the population boundary or mixing conclusions across evidence types.^20,21^ These challenges become particularly important when evidence is absent, indirect or discordant across study designs.

Multi-agent systems (MAS)^22–24^ provide a natural way to organize evidence synthesis around these distinct tasks.^25–27^ In a multi-agent workflow, specialized agents can retrieve published literature, generate question-specific RWE by constructing and analyzing cohorts from underlying real-world data, query biomedical knowledge graphs, perform statistical appraisal and synthesis and verify claims before a final report is produced.^22^ This separation is useful because each evidence source has different strengths and failure modes. Published studies may be affected by selective reporting or narrow eligibility criteria; generated real-world analyses may be affected by confounding, measurement error and incomplete ascertainment; and knowledge graphs may be affected by incomplete or uneven curation. A coordinated system can therefore preserve these evidence streams separately, evaluate their relevance, uncertainty and applicability and synthesize them without treating agreement as proof of validity or forcing discordant sources into an unqualified consensus.

Here we present Multi-Agent Evidence Synthesis (MES), a multi-agent framework that retrieves published evidence, generates question-specific RWE from real-world clinical data and incorporates structured biomedical knowledge while preserving the provenance and limitations of each evidence source. MES comprises six specialized agents: a Supervisor for query decomposition and orchestration; a LiteratureMiner for biomedical literature and trial evidence; an RWD-Analyst that translates clinical questions into computable phenotypes, constructs real-world cohorts and executes corresponding analyses; a knowledge graph (KG)-Specialist for structured biomedical knowledge retrieval and mechanistic context; a Statistician for evidence appraisal, quantitative synthesis when study designs and estimands are sufficiently compatible and cross-source divergence assessment; and a SafeChecker for claim-level verification. The Supervisor structures user questions using evidence-based medicine frameworks, including PICO^28,29^ and the test–target condition–reference standard (TTR) framework, asks for clarification when key elements are missing and triggers additional analyses when evidence gaps or disagreements are detected. SafeChecker decomposes the draft synthesis into individual claims, traces each claim to its source evidence and checks consistency across agent outputs.

We evaluated MES in two complementary ways. First, we illustrate its behavior in two clinical use cases representing different evidence conditions: an individualized Alzheimer disease question for which matching published evidence was absent and a septic shock treatment question for which directly relevant randomized evidence was available but inconsistent. These examples test whether MES can identify evidence gaps, generate question-specific real-world evidence when appropriate, preserve distinctions between observational and randomized evidence, report unresolved uncertainty and avoid unsupported recommendations. Second, we evaluate MES across 144 clinical queries spanning six evidence-based medicine categories, using automated metrics and manual expert assessment across four LLM backbones. We further examine cross-source integration, evidence-divergence identification and claim-level evidence-support screening using the CliniFact benchmark. Together, these analyses test whether MES can generate structured evidence reports that are source-grounded, clinically interpretable and explicit about uncertainty and evidentiary boundaries. MES is not intended to replace systematic reviews or expert judgement, nor does the present evaluation establish the clinical utility or transportability of MES-generated RWE. Rather, MES is designed as an auditable framework for generating, organizing and contextualizing heterogeneous evidence while identifying where evidence is sufficient, conflicting or absent.

## Methods

### Ethics Statement

This study was approved by the Institutional Review Board of Weill Cornell Medicine with protocol number 21-07023759. All EHR data used in this study were fully deidentified; therefore, informed consent was not required.

### Data

**MIMIC-IV.**^30^ We used the Medical Information Mart for Intensive Care (MIMIC-IV), a critical-care EHR resource containing records from more than 299,000 patients treated at Beth Israel Deaconess Medical Center in Boston, Massachusetts, from 2008 to 2019. MIMIC-IV provides time-resolved information on physiology, laboratory measurements, interventions and other encounter-level clinical variables during intensive-care encounters. In this study, MIMIC-IV supported evidence-synthesis tasks involving acute and rapidly evolving conditions, including septic shock, acute heart failure and acute kidney injury. These tasks require temporally granular data to characterize patient status, treatment exposure and outcomes.

**INSIGHT.**^31^ We further used data from the INSIGHT Clinical Research Network (CRN), a large longitudinal EHR network from five health systems in the greater New York City area, covering the period from January 2007 through December 2023. In contrast to ICU-focused datasets, INSIGHT provides longitudinal clinical histories across repeated healthcare encounters, making it well suited for studying chronic disease progression and long-term treatment patterns.

### MES Architecture Overview

MES is a modular multi-agent framework for medical evidence synthesis from three evidence classes: published biomedical literature and trial registries, real-world clinical data from EHRs and structured biomedical knowledge graphs. The framework coordinates six agents: a Supervisor, LiteratureMiner, RWD-Analyst, KG-Specialist, Statistician and SafeChecker (Fig. 1). Each agent has a defined role and returns structured outputs with provenance metadata.

**Figure 1.**
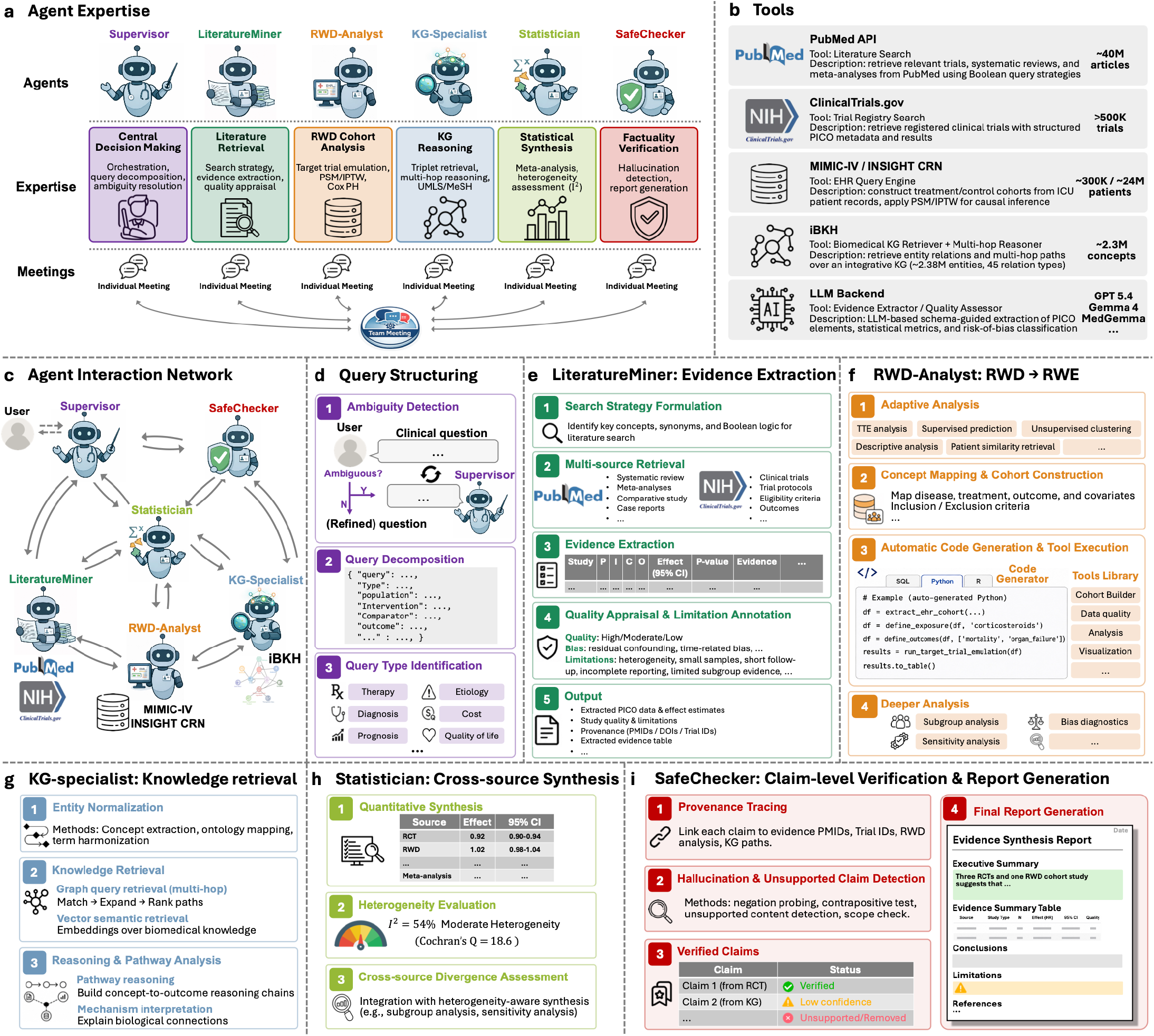
Overall framework of MES. (a) Agent expertise. MES is organized around six specialized agents. The Supervisor performs query decomposition, workflow orchestration and ambiguity resolution. LiteratureMiner retrieves and extracts evidence from biomedical literature. RWD-Analyst constructs real-world cohorts and performs causal or predictive analyses. KG-Specialist retrieves and reasons over biomedical knowledge graphs. Statistician appraises quantitative evidence, performs synthesis when evidence is sufficiently comparable, assesses heterogeneity and characterizes cross-source divergence. SafeChecker screens claim-level evidentiary support and consistency, flags unsupported claims and supports final report generation. **(b) Tools.** MES connects agents to PubMed, ClinicalTrials.gov, MIMIC-IV, INSIGHT CRN, iBKH and LLM backends for evidence extraction, quality assessment and structured reasoning. **(c) Agent interaction network.** The Supervisor coordinates communication between the user and specialist agents. LiteratureMiner, RWD-Analyst and KG-Specialist collect complementary evidence from literature, RWD and KG sources. Statistician appraises and integrates quantitative findings, while SafeChecker verifies claims and checks consistency before final synthesis. **(d) Query structuring.** The Supervisor detects ambiguity, refines the user question, decomposes it into structured elements and identifies the clinical question type, including therapy, diagnosis, prognosis, etiology, cost or quality of life. **(e) LiteratureMiner**. This agent formulates search strategies, retrieves evidence from PubMed and ClinicalTrials.gov, extracts study characteristics and effect estimates, appraises study quality, annotates limitations and outputs structured evidence summaries. **(f) RWD-Analyst.** This agent converts real-world data into real-world evidence through adaptive analysis selection, concept mapping, cohort construction, automatic code generation, tool execution, subgroup analysis, sensitivity analysis and bias diagnostics. **(g) KG-Specialist.** This agent normalizes biomedical entities, retrieves graph evidence and performs multi-hop pathway reasoning to support mechanistic interpretation. **(h) Statistician.** This agent harmonizes evidence across sources, performs quantitative synthesis when appropriate, evaluates heterogeneity using statistics such as I² and Cochran’s Q, and identifies and characterizes cross-source disagreement. **(i) SafeChecker.** This agent traces provenance, detects hallucinated or unsupported claims, labels verified and low-confidence claims and produces the final evidence synthesis report with conclusions, limitations and references.

The Supervisor receives a natural-language clinical or research question and converts it into a structured task representation. It identifies the clinical question type, detects missing or ambiguous elements, selects the evidence modules required for the task and coordinates inter-agent communication. LiteratureMiner retrieves and extracts evidence from biomedical literature and trial registries. RWD-Analyst constructs EHR cohorts and performs causal, predictive, descriptive, clustering or patient-similarity analyses. KG-Specialist retrieves ontology-grounded relations and graph paths from biomedical knowledge graphs. Statistician harmonizes and integrates quantitative findings across evidence sources. SafeChecker verifies claim support and consistency before final report generation.

The synthesis workflow is iterative. Initial query decomposition may trigger clarification requests when clinically important elements are missing. Specialist agents then retrieve or analyse source-specific evidence, either in parallel or through targeted follow-up calls. When evidence is absent, incomplete or discordant, agents can request additional retrieval, stratified analysis or verification. The process stops when the decomposed query has been addressed, when unresolved uncertainty has been explicitly recorded or when the available sources have been exhausted. Throughout the workflow, MES records the source, agent and processing step that support each final claim.

## Agents

Each agent in MES is designed to perform a specific set of tasks as summarized below:

- *<u>Supervisor</u>*: The central coordinator that decomposes user queries, maintains dialogue context, resolves ambiguity, triggers clarification or fallback routines, coordinates specialist agents and integrates their outputs into a final structured synthesis while maintaining system-level coherence.
- *<u>LiteratureMiner</u>*: Responsible for retrieving and synthesizing evidence from literature sources, such as PubMed, ClinicalTrials.gov, and systematic reviews. Extracts population-specific findings, intervention-outcome relationships, and study metadata with provenance-aware citation support.
- *<u>RWD-Analyst</u>*: Handles real-world evidence generation from real-world clinical data and causal inference modeling. Constructs treatment/control cohorts, estimates comparative effectiveness or risk, and provides adjusted statistical metrics (e.g., hazard ratios, confidence intervals). Supports subgroup analysis (e.g., age, comorbidity strata) and sensitivity analysis to assess heterogeneity of treatment effects and robustness of findings.
- *<u>KG-Specialist</u>*: Interfaces with biomedical knowledge graphs (KGs) to extract structured, ontology-grounded information (e.g., drug-disease relationships, contraindications). Performs multi-hop reasoning, constraint checking, and supports concept normalization to bridge unstructured and structured evidence.
- *<u>Statistician</u>*: Performs statistical synthesis and integration across multiple agents, including meta-analysis, subgroup analysis, and heterogeneity evaluation. Quantifies uncertainty from effect estimates and confidence intervals, assesses whether evidence is sufficiently comparable for quantitative synthesis, identifies divergence across evidence sources and outputs pooled estimates or statistical visualizations.
- *<u>SafeChecker</u>*: Acts as an evidence-support screening and quality-control layer. It assesses claim-level support relative to supplied evidence, traces provenance, checks inter-agent consistency and flags unsupported or low-confidence outputs. It annotates each final claim with a screening status, confidence score and evidence-traceability metadata; these annotations are not guarantees of factual or clinical correctness.

### MES for Evidence Synthesis

#### Query decomposition and agent routing

Given a natural-language clinical question, the Supervisor converts the query into a structured representation that guides downstream agent actions. It assesses whether the input is complete enough for evidence retrieval and analysis. When important components are missing, the Supervisor initiates clarification before activating specialist agents.

The Supervisor autonomously evaluates the nature of each clinical question and selects the most appropriate evidence-based medicine framework for decomposition. Rather than applying fixed rules, the agent dynamically determines the question categories including but not limited to Therapy, Diagnosis, Prognosis, Etiology/Harm, Cost-effectiveness, and Quality of life based on its analysis of the query content and clinical context. When the agent identifies a therapy question, it adopts the PICO framework to extract Population, Intervention, Comparator, and Outcome components. For diagnostic inquiries, the agent applies the TTR schema to identify the Test, Target condition, and Reference standard. The agent adapts its decomposition approach based on the specific characteristics of each query, extracting relevant components and validating their completeness. For instance, when processing “metformin’s effect on cognitive decline in elderly diabetic patients,” the agent recognizes this as a therapy question and systematically extracts the population, intervention, and outcome while identifying the need to clarify the comparator.

When the Supervisor detects incomplete or ambiguous components during decomposition, it autonomously initiates clarification dialogue with the user.^26^ The agent evaluates each extracted component for clinical specificity, identifies gaps that could affect evidence retrieval quality, and makes contextual decisions about when clarification is necessary based on the query completeness and downstream evidence synthesis requirements. When detecting an incomplete therapy question lacking outcome specification, the Supervisor generates contextually appropriate clarification prompts such as: “What clinical outcome are you interested in evaluating? For example, mortality, disease progression, quality of life measures, or laboratory parameters?” The clarification process operates iteratively, with the agent maintaining dialogue context and incorporating user responses to refine the query representation. The agent autonomously determines when sufficient information has been obtained to proceed with evidence synthesis, balancing thoroughness with user burden. This adaptive approach allows the agent to handle varying levels of query complexity and user expertise while ensuring that downstream agents receive adequately structured information for their tasks.

Following query analysis and clarification, the Supervisor evaluates the clinical question and recommends which specialist agents to engage based on the evidence requirements. It identifies the agents best suited to address the decomposed question and initiates the multi-agent evidence synthesis workflow. These routing decisions determine which analyses are prioritized, but they do not assign evidentiary weight; evidence from all activated agents is subsequently appraised by the Statistician according to study design, relevance, uncertainty and applicability.

### Retrieve and Summarize Evidence from Literature

Upon receiving the structured query, the Supervisor activates the LiteratureMiner agent to conduct a systematic search and analysis of published biomedical literature from sources like PubMed and ClinicalTrials.gov. This process is executed in a multi-step pipeline.

First, the agent formulates a search strategy. It analyzes the decomposed query to determine the optimal data source; for example, questions about established interventions are directed primarily to PubMed, while those concerning novel therapies may prioritize ClinicalTrials.gov. The agent then generates search strings, including Boolean operators and structured syntax where appropriate, to retrieve a focused set of relevant clinical trial records and publication abstracts. Next, the Evidence Extractor module processes each retrieved document to convert unstructured text into a structured format. Using a schema-guided approach, the agent prompts an LLM to populate a predefined JSON object by identifying and extracting the core PICO elements and any quantitative statistical metrics, such as odds ratios (OR), hazard ratios (HR), and their corresponding confidence intervals and p-values. Following extraction, each piece of evidence undergoes critical appraisal. The Study Quality Assessor evaluates the methodological rigor of the source study. It assesses key details such as study design (e.g., randomized controlled trial, retrospective cohort), population characteristics and the clarity of the reported endpoints to assign a provisional risk-of-bias screening label (”Low”, “Moderate”, or “High”) with a concise justification. This label is used for internal evidence appraisal and is not a substitute for a formal study-specific risk-of-bias assessment. In parallel, the Limitation Annotator performs a targeted scan of the source text, specifically searching for keywords and phrases that denote scientific caveats (e.g., “small sample size,” “observational nature,” “results may not be generalizable”). These limitations are extracted verbatim to ensure their original context is preserved. Finally, the Evidence Aggregator consolidates these layers of information (the structured PICO data, statistical metrics, quality assessment, and annotated limitations) for each document. This enriched data is compiled into a summary table, and a narrative synthesis is generated to provide a coherent overview of the findings from the literature. This structured output is then passed back to the Supervisor for subsequent verification and cross-synthesis with evidence from other agents.

### Extract Real-world Evidence from Real-world Data

To complement published evidence, the RWD-Analyst dynamically interrogates real-world clinical data from MIMIC-IV or the INSIGHT Clinical Research Network, depending on the clinical question and required temporal context. MIMIC-IV supports acute and critical-care analyses requiring temporally granular encounter data, whereas INSIGHT supports longitudinal analyses requiring clinical histories across repeated healthcare encounters. A preprocessing module first extracts medical concepts from the free-text clinical query and maps them to standard terminologies, such as ICD-10 and LOINC codes, to construct an analysis-ready dataset. Rather than relying on a single analytical approach, the system employs a language model-driven routing mechanism to classify the analytical intent and direct the data to one of five automated pipelines: target trial emulation, supervised prediction, descriptive analysis, unsupervised clustering or patient similarity retrieval.

For queries requiring causal inference, the agent executes a comprehensive target trial emulation protocol. It translates the clinical criteria into a formal study design, extracting explicit treatment and comparator cohorts. When treatment assignment is time dependent and would otherwise create immortal time bias, the pipeline applies a clone-censor-weight methodology.^32^ The agent addresses measured baseline confounding through covariate balancing, applying either propensity score matching or inverse probability of treatment weighting, and assesses post-adjustment balance using standardized mean differences (SMDs^33^). These procedures do not eliminate residual or unmeasured confounding. The treatment effect is then estimated via models such as Cox Proportional Hazards or Random Survival Forests.^34^ When clinically relevant subgroup questions are prespecified and sufficient data are available, the agent can perform stratified analyses to assess potential heterogeneity of treatment effects and generate hypotheses for further evaluation. Beyond causal inference, the agent functions as an automated analytical suite to address diverse clinical inquiries. For prognostic questions, a supervised learning pipeline trains regularized regression models, evaluates performance using area under the receiver operating characteristic curve metrics, and extracts the ten most influential predictive features. Exploratory queries trigger an unsupervised clustering pipeline that applies K-means algorithms, dynamically optimizing the number of clusters via silhouette scores to discover latent patient phenotypes. For cohort characterization, a descriptive analysis module computes comprehensive summary statistics to generate a standardized baseline profile. Case-based queries activate a patient retrieval pipeline that computes Euclidean distances across scaled feature spaces to identify historically similar patient records, providing empirical context for specific clinical presentations. Regardless of the executed pipeline, the resulting statistical output is packaged into a standardized format and treated as an independent empirical study within the broader system architecture. This output is routed directly to the Statistician agent.

### Extract Structured Knowledge from Biomedical Knowledge Graph

To complement empirical findings from real-world data and literature, MES leverages the KG-Specialist agent to provide ontology-grounded insights from biomedical knowledge graphs (KGs). By retrieving curated relationships between biomedical entities, such as drugs, diseases, genes, and pathways, this agent introduces mechanistic, regulatory, and hierarchical evidence that strengthens the interpretability and consistency of the overall synthesis.

The KG-Specialist begins by retrieving triplets relevant to the input question from the knowledge graphs, which support both structured and vector-based retrieval methods. The agent selects between the two modes based on the input question, using structured retrieval when entities can be reliably mapped to KG nodes and vector-based retrieval otherwise. In the structured retrieval mode, the agent maps entities in the input question to KG nodes using UMLS CUIs or MeSH IDs^35^, then generates queries in graph query languages including Cypher^36^ and SPARQL^37^ for execution. Generating queries for multi-hop reasoning over KGs is also available in this mode for querying indirect relationships between entities. In the vector-based retrieval mode, a vector database is pre-built for each knowledge graph by embedding all its triplets into feature vectors using embeddinggemma-300m-medical^38,39^, a Gemma 3-based model adapted for biomedical/clinical text retrieval. Given an input question, the agent embeds it into a feature vector with the same model and retrieves the top-k triplets whose vectors have the highest cosine similarity to the question vector. After obtaining the retrieved triples and multi-hop reasoning paths, the KG-Specialist quantifies their reliability based on (1) the quality of the original source (”Low”, “Moderate”, or “High”), and (2) the consistency across the retrieved triples and reasoning paths.

### Perform Statistical Synthesis and Integration

After the specialist agents have gathered evidence, the Statistician agent appraises the quantitative evidence and determines whether cross-study synthesis is appropriate. It considers study design, target population, intervention and comparator definitions, outcome measures, effect estimands, precision and potential sources of bias when assessing comparability across evidence sources. The agent does not assign a fixed numerical weight to different evidence classes. Instead, evidence that meets these comparability criteria undergoes quantitative synthesis, whereas evidence that differs materially in design, estimand or population is retained separately and evaluated for agreement or disagreement.

The process begins with normalization of incoming evidence from the LiteratureMiner, RWD-Analyst and KG-Specialist. For quantitative evidence, the Statistician extracts effect estimates, confidence intervals, sample sizes and other relevant statistical information and converts them into a standardized representation for comparison and, when appropriate, meta-analysis. When quantitative parameters are incomplete, the agent derives missing quantities where feasible, such as calculating standard errors from reported confidence intervals using established statistical relationships. Evidence is categorized by effect-measure type and study design because different estimands and designs may require different analytical treatment. Evidence lacking essential quantitative components is excluded from pooled analyses but remains available for qualitative and contextual interpretation.

When multiple quantitative studies address the same clinical question, the Statistician evaluates whether quantitative pooling is appropriate. Pooling requires sufficient compatibility in study design, target population, intervention and comparator definitions, outcome measurement and effect estimand. Evidence arising from materially different study designs, particularly randomized trials and newly generated observational RWE, is ordinarily summarized separately rather than combined into a single pooled estimate unless their analytical targets and assumptions are sufficiently aligned. For studies considered suitable for pooling, the agent aggregates compatible effect estimates, typically using a random-effects model to account for within-study sampling error and between-study variability. When only a single valid study is available for an effect measure, the individual estimate is presented without pooling. The agent also generates structured data for standard meta-analytic visualizations, including forest plots showing individual and pooled estimates when applicable.

For studies that are pooled, the Statistician quantifies statistical heterogeneity using the I² statistic.^40,41^ I² describes inconsistency among effect estimates but does not identify its cause or determine which evidence source is more credible. The agent therefore considers potential clinical and methodological explanations for observed heterogeneity, including differences in populations, intervention definitions, outcome ascertainment and study design. When sufficient comparable data are available, subgroup analyses may be used to explore prespecified sources of heterogeneity. These analyses are treated as exploratory assessments of effect variation and do not resolve residual confounding or other sources of bias in observational evidence.

When evidence sources yield conflicting findings, the Statistician treats the discrepancy as cross-source disagreement rather than assuming that it can be resolved statistically. The agent compares the underlying study designs, populations, eligibility criteria, intervention and comparator definitions, outcome ascertainment, follow-up periods and effect estimands to identify potential explanations for the discrepancy. Differences in risk of bias and potential confounding are also considered when interpreting observational evidence. For generated RWE, this appraisal incorporates the available cohort and phenotype definitions, treatment and outcome specifications, covariate-balance diagnostics and identified data limitations; residual or unmeasured confounding and limited transportability are retained as unresolved limitations rather than treated as statistically resolved. For published studies, the Statistician uses the study-quality and limitation annotations supplied by the LiteratureMiner. When the evidence sources remain in conflict after these comparisons, their estimates are reported separately rather than combined or ranked solely on the basis of statistical significance. The final synthesis preserves the disagreement, describes plausible methodological or clinical explanations and explicitly reports the remaining uncertainty.

### Verify Factuality and Control Quality

The final stage before the Supervisor synthesizes the report is a critical quality control process managed by the SafeChecker agent. This agent acts as the system’s verifier, assessing whether generated claims are attributable to and consistent with the supplied evidence before they are presented to the user.

The core function of the SafeChecker is verification on the evidence summary generated by the Statistician based on all evidence used to generate such evidence summary. To begin, the SafeChecker decomposes the summary into individual claims. A claim is a concise, verifiable statement that expresses a specific conclusion or relationship derived from the evidence, serving as the basic unit for subsequent validation. Then, the SafeChecker traces each claim back to the originating evidence, whether it is a passage from a publication, a result from a real-world data analysis, or from a knowledge graph. This process assesses whether each claim is directly supported by its cited source and flags potential misinterpretation or exaggeration of findings. First, the Hallucination Detector module will verify if the claim can be linked to any provided evidence by contrapositive / negation probing. Claims flagged by the Hallucination Detector^42^ are recommended for removal or further review.

In addition to verifying individual claims, the Consistency Checker module evaluates the logical coherence across the entire evidence set. It identifies contradictions, such as when findings from literature conflict with results from the RWD analysis for the same population and outcome. The agent also leverages biomedical constraints, potentially retrieved by the KG-Specialist, to ensure that synthesized claims do not violate known medical principles. The SafeChecker annotates each piece of evidence with a verifiability status and a confidence score based on the quality and agreement of the underlying sources. Low-confidence or unsupported claims are flagged and reported to the Supervisor, which may trigger a fallback routine, a request for clarification from another agent, or the exclusion of the claim from the final report.

### Final Report Synthesis and Iterative Refinement

The Supervisor integrates verified outputs into a structured evidence report. The report includes a title, executive summary, source-specific evidence sections, quantitative result tables, visualizations when available, limitations and citations. The organization of the report depends on the clinical question. Therapy and comparative-effectiveness questions emphasize clinical effectiveness, comparative outcomes and safety. Diagnostic questions emphasize test performance, reference standards and population-specific accuracy. Prognostic and health-services questions emphasize outcome definition, applicability and decision boundaries.

Before finalization, the Supervisor checks report completeness against PRISMA-lite-style criteria.^43,44^ These checks assess whether the clinical question is stated, evidence sources are described, quantitative results are reported with sufficient detail, limitations are presented and conclusions remain balanced. The Supervisor also compares the draft report with the decomposed query to determine whether all required elements have been addressed.

If the report remains incomplete, the Supervisor launches targeted follow-up work rather than repeating the full pipeline. For example, if literature evidence exists in younger adults but not older adults, the Supervisor can request geriatric-focused retrieval from LiteratureMiner and age-stratified analysis from RWD-Analyst. Each refinement cycle is followed by SafeChecker verification. The workflow terminates when the query is answered, when unresolved uncertainty is documented or when available evidence sources are exhausted.

### Evaluation Query Set

The evaluation set comprised 144 clinical queries with balanced coverage across six evidence-based medicine (EBM) categories and eight clinical domains. The six EBM categories were clinical effectiveness, comparative effectiveness, diagnostic accuracy, prognosis or risk, safety and adverse events, and cost-effectiveness. The eight clinical domains were cardiology, oncology, endocrinology, infectious diseases, psychiatry, emergency medicine, pediatrics, and geriatrics. Each EBM category– domain combination included three queries, yielding 144 queries in total.

### Theoretical Analysis

The theoretical analysis formalizes five design choices in MES: decomposition into specialist agents, heterogeneous-source integration, claim verification, iterative refinement and generalization under distribution shift.^45,46^ These results are conditional statements under explicitly specified mathematical and algorithmic models. They clarify sufficient conditions for selected design properties but do not establish that those conditions hold empirically or constitute evidence of clinical validity. The estimation results in Supplementary Notes 2–3 additionally require source outputs to target the same estimand on a common scale; they do not justify numerically averaging incommensurable trial effects, observational associations and mechanistic graph relations. Full definitions, assumptions, theorem statements and proofs are provided in Supplementary Notes 2–6.

### A multi-agent design can reduce estimation risk relative to a single-agent design

A single end-to-end agent must resolve retrieval, analysis and synthesis tasks within a shared context. By contrast, MES assigns each evidence stream to a dedicated specialist agent and subsequently integrates their outputs. We therefore ask whether this decomposition provides a formal advantage beyond its practical convenience. To ensure a conservative comparison, we grant the monolithic baseline the same source-specific bias as the specialist agents and allow it to choose its own optimal internal mixture. We analyze specialization under an explicit joint interference model. Let 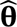 denote the vector of specialist source estimates and let the corresponding monolithic sub-estimates be 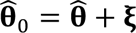. The sufficient joint condition 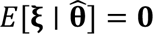 preserves the bias vector and eliminates all specialist–interference cross-covariances, yielding 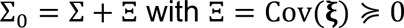. This is a conditional model of capacity interference; componentwise mean-zero noise alone would not be sufficient for the matrix ordering. Let *R*_MA_ and *R*_SA_ denote the mean-squared-error risks of the optimally aggregated multi-agent and monolithic estimators, respectively. Supplementary Theorem 2.1 establishes that

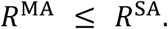

The inequality is strict when the monolithic optimal weighting direction has positive interference variance, 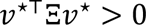. In the unbiased, mutually uncorrelated positive-variance special case, the optimal aggregation weights are inverse-variance weights and

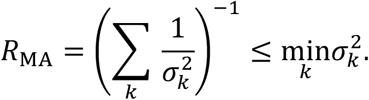

With *K* ≥ 2, this special case yields a strict diversification gain. The conclusion does not follow merely from the informal description that sources are “non-redundant.”

This analysis motivates studying the separation of literature, real-world data and knowledge-graph retrieval into specialist agents. Under the joint PSD covariance-inflation model, specialization is non-inferior in oracle mean-squared-error risk and is strictly beneficial when the monolithic optimal weighting direction has positive interference variance. “Complementary information” alone is not the formal condition, and the theorem does not establish that the covariance-inflation model holds for the evaluated LLMs.

### Integrating heterogeneous evidence sources can reduce systematic bias

Literature, real-world data and knowledge graphs are subject to distinct sources of systematic error, including publication bias, residual confounding and incomplete curation. It is therefore not self-evident that integrating multiple imperfect evidence streams is preferable to relying on the strongest individual source. We ask whether convex fusion can reduce systematic bias and identify a sufficient condition under which the improvement is strict. Let *ρ*\* = min_w_ ∥ ∑*_s_ w_s_ b_s_* ∥ denote the smallest achievable bias under convex fusion, where *b_s_* is the bias associated with source. Let *m* = min*_s_* ∥ *b_s_* ∥ denote the bias of the least-biased individual source. Supplementary Note 3 (Theorem 3.1) establishes that *ρ*\* ≤ *m*. Thus, the population-level bias frontier achieved by optimally weighted fusion is never more biased than the best individual source under the stated model. This is an oracle statement because the bias-optimal weights depend on the unknown source biases. Moreover, the inequality is strict whenever there exists a source *j* satisfying the directional-diversity condition

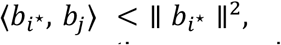

Geometrically, strict improvement requires more than non-collinearity. For a least-biased source *i*\*, at least one source *j* must satisfy

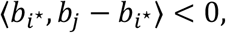

so that moving from the vertex *b_i_*★ toward *b*_j_ decreases the squared bias norm to first order. Sources with distinct error mechanisms need not satisfy this directional condition; it is an explicit population-level hypothesis.

Supplementary Note 3 also defines an oracle risk frontier. Because every single-source vertex is feasible, the minimum risk over convex mixtures cannot exceed the best single-source risk; strict improvement requires the directional derivative condition stated in Theorem 3.4. The general optimizer depends on unknown source biases and cross-covariances and is therefore not directly computable. The unbiased, mutually uncorrelated special case is different: inverse-variance weighting is computable from source variances and strictly improves risk when at least two positive-variance sources are available. MES uses uncertainty, study quality, provenance and cross-source consistency as signals for source appraisal and synthesis, but the formal result does not certify the deployed decision rules.

This result motivates studying the integration of literature, real-world data and knowledge-graph evidence within MES. These sources may have different error mechanisms, but distinct provenance alone does not establish the required population-level bias directions. When the displayed directional-diversity condition holds, an oracle convex mixture can be less biased than every individual evidence stream. MES does not observe the true bias vectors; its uncertainty, quality and provenance signals are practical proxies rather than proof that the oracle frontier is attained. The Statistician therefore reports uncertainty and does not treat consensus as proof that bias has been eliminated.

### SafeChecker provides finite-sample control of false-claim acceptance

Verification is useful only if it provides an explicit guarantee on the rate at which false claims enter the final report. We therefore analyze a calibrated version of SafeChecker in which a nonconformity score is calibrated using a reference set of known-false claims. The objective is to determine what can be guaranteed without imposing a parametric model on the score distribution. For a user-specified tolerance level *α* and a calibration set containing *N* known-false claims, Supplementary Note 4 (Theorem 4.1) establishes the following finite-sample, distribution-free bound:

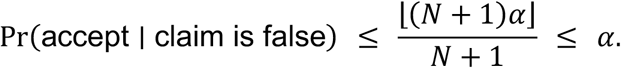

Accordingly, if *H_i_* denotes the event that a false claim *i* is accepted into the report and *T_i_* = 0 indicates that it is false, then

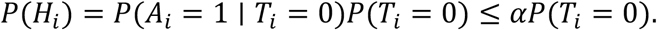

Thus, SafeChecker does not eliminate false claims but bounds the probability that they pass the calibrated screen under the stated assumptions. For multiple claims, report-level control additionally requires null super-uniformity conditional on the report’s truth configuration. Conditional mutual independence of the full *p*-value vector, or PRDS on the true-null coordinates, permits Benjamini– Hochberg control; because claims in a report may share evidence and generation context, the Benjamini–Yekutieli correction is the guaranteed option under arbitrary conditional dependence (Theorem 4.4).^47^

This analysis characterizes SafeChecker as a calibrated screening mechanism rather than an oracle. The tolerance level *α* determines the false-acceptance ceiling under the stated exchangeability assumptions. Lower values impose more conservative screening, potentially at the cost of rejecting or abstaining on more valid claims. The identity separates two quantities: the upstream probability that a false claim is generated and the conditional probability that such a claim passes screening. Theorem 4.1 controls only the second quantity. Reducing the upstream false-claim probability is a system objective evaluated empirically here, not a consequence of Supplementary Notes 2–3.

### Conditional termination and residual upper bounds for iterative refinement

An iterative synthesis procedure is useful only if it stops under explicit progress conditions and avoids persistent unresolved or contradictory states. We define a non-negative synthesis residual *V_t_* that aggregates unresolved uncertainty, flagged contradictions and ungrounded claims at refinement round *t*. Finite corpora and a finite aspect list do not by themselves prevent repeated or stalled updates. Supplementary Theorem 5.1 therefore states a conditional sufficient-condition guarantee. For a fixed query, let A be a finite set of admissible refinement opportunities and let *Q_t_* ⊆ A be the completed opportunities after round *t*. Suppose the process stops when the residual tolerance is met or when *Q_t_* = A ; otherwise, completed opportunities are never repeated and every non-terminal round completes at least one previously unused opportunity. Then

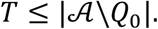

At termination, the process has either met the residual tolerance or exhausted the admissible refinement opportunities. The theorem does not assert that the current implementation unconditionally satisfies these assumptions, and termination does not certify that uncertainty has vanished or that the report is clinically correct. A separate result applies if the residual satisfies

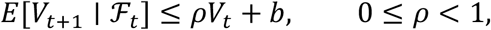

where ℱ*_t_* is the information available through round *t* and *b* ≥ 0 is an additive drift allowance. Supplementary Theorem 5.2 then gives

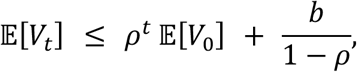

The transient term in this upper bound decays geometrically, and

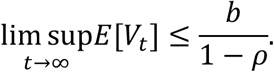

For *b* > 0, this is an upper-envelope statement: it does not imply that *E*[*V_t_*] converges, that *V_t_* approaches a positive “noise floor” or that *b*/(1 − *ρ*) is a lower bound. When *b* = 0, the theorem additionally gives *E*[*V_t_*] → 0 geometrically and *V_t_* → 0 almost surely.

The finite-termination guarantee applies only under the finite-progress conditions formalized in Theorem 5.1: a finite admissible refinement set, no repetition and progress or stopping at every non-terminal iteration. Under the separate drift condition of Theorem 5.2, the expected residual is bounded by a geometrically decaying transient plus an additive asymptotic envelope. This controls a residual upper bound and does not, for *b* > 0, establish convergence of the actual residual sequence. A low-residual or refinement-exhausted state is therefore a stopping condition, not a guarantee that uncertainty has been eliminated.

### Generalization to target queries under distribution shift

Supplementary Note 6 studies an idealized supervised-learning setting in which a synthesis function is trained from independent labeled examples supplied by multiple source distributions. This is a conditional learning-theory model, not a direct probabilistic description of arbitrary publications, EHR analyses and graph relations produced by the deployed workflow. For weights *w* fixed independently of the observed source samples, Supplementary Theorem 6.B gives, with probability at least 1 − *δ* and uniformly over

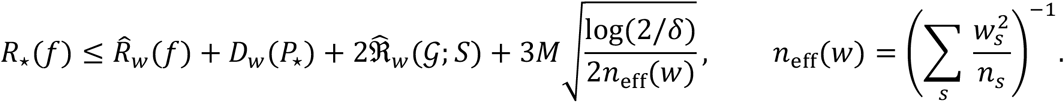

The hatted Rademacher term is empirical. The discrepancy *D*_w_(*P*_*_) is a population quantity unless it is estimated or upper-bounded using suitable target data. Thus, the result is a conditional target-risk bound, not automatically a computable certificate for the deployed MES workflow. The bound displays a trade-off among empirical fit, source–target discrepancy, class complexity and effective sample size. For a prespecified finite library of candidate weights, a union bound permits valid post-selection of the smallest full certificate (Corollary 6.C.1). The selected candidate may be a vertex, boundary point or interior mixture. Unrestricted query-adaptive optimization on the same data is not covered without an independent tuning split or a theorem uniform over the continuous weight simplex.

MES heuristically combines literature, real-world data and knowledge-graph evidence using relevance, provenance and uncertainty signals. The revised bound motivates factors that source appraisal and synthesis should consider, but it certifies only fixed weights or selection from a prespecified finite library under the stated sampling and discrepancy assumptions. It does not establish that the deployed synthesis procedure minimizes deployment risk.

### User Interface

MES provides an interactive web interface that allows clinicians and researchers to initiate, monitor, and inspect multi-agent evidence synthesis in a transparent manner. The interface is designed around a question-driven workflow, in which users enter a natural-language clinical or research question and select the evidence sources to be activated, including published literature, knowledge graphs, and real-world clinical data. To reduce the burden of formal query formulation, the interface also provides task-oriented templates corresponding to common evidence-based medicine question types, such as therapy, etiology/harm, prognosis, and cost-effectiveness. This design enables users to begin with an informal clinical question while allowing the system to convert the query into structured components for downstream evidence retrieval, real-world data analysis, knowledge graph reasoning, statistical synthesis, and safety verification.

After a query is submitted, the interface displays the synthesis process as a structured investigation workspace. The results panel summarizes the original research question, retrieved evidence from the activated sources, key findings, and the final synthesized answer. Each evidence source is presented separately for inspection, allowing users to examine how literature-derived findings, real-world data outputs, and knowledge graph evidence contribute to the final answer. The synthesized response retains source-grounded citations and evidence provenance, enabling users to trace individual conclusions back to the supporting evidence. This modular presentation supports source-level transparency and prevents the final synthesis from appearing as an opaque language-model response.

The interface further provides specialized inspection panels for each agent. The LiteratureMiner workspace presents retrieved studies, study details, extracted PICO elements, and reported quantitative effect estimates, including subgroup-specific metrics when available. The RWD-Analyst workspace supports target trial specification, cohort definition, inclusion and exclusion criteria, treatment definition, cohort construction, outcome summaries, and covariate balance assessment based on real-world clinical data. The KG-Specialist workspace enables users to inspect biomedical entities, relationships, source databases, and graph-based connections among drugs, diseases, and related clinical concepts. Together, these panels allow users to trace how MES transforms a clinical question into a multi-source evidence report, while preserving interpretability at each step of agent reasoning and evidence synthesis. This UI design is aligned with the MES architecture, which integrates literature, real-world data, knowledge graphs, statistical synthesis, and factual verification through specialized agents.

**Figure 2.**
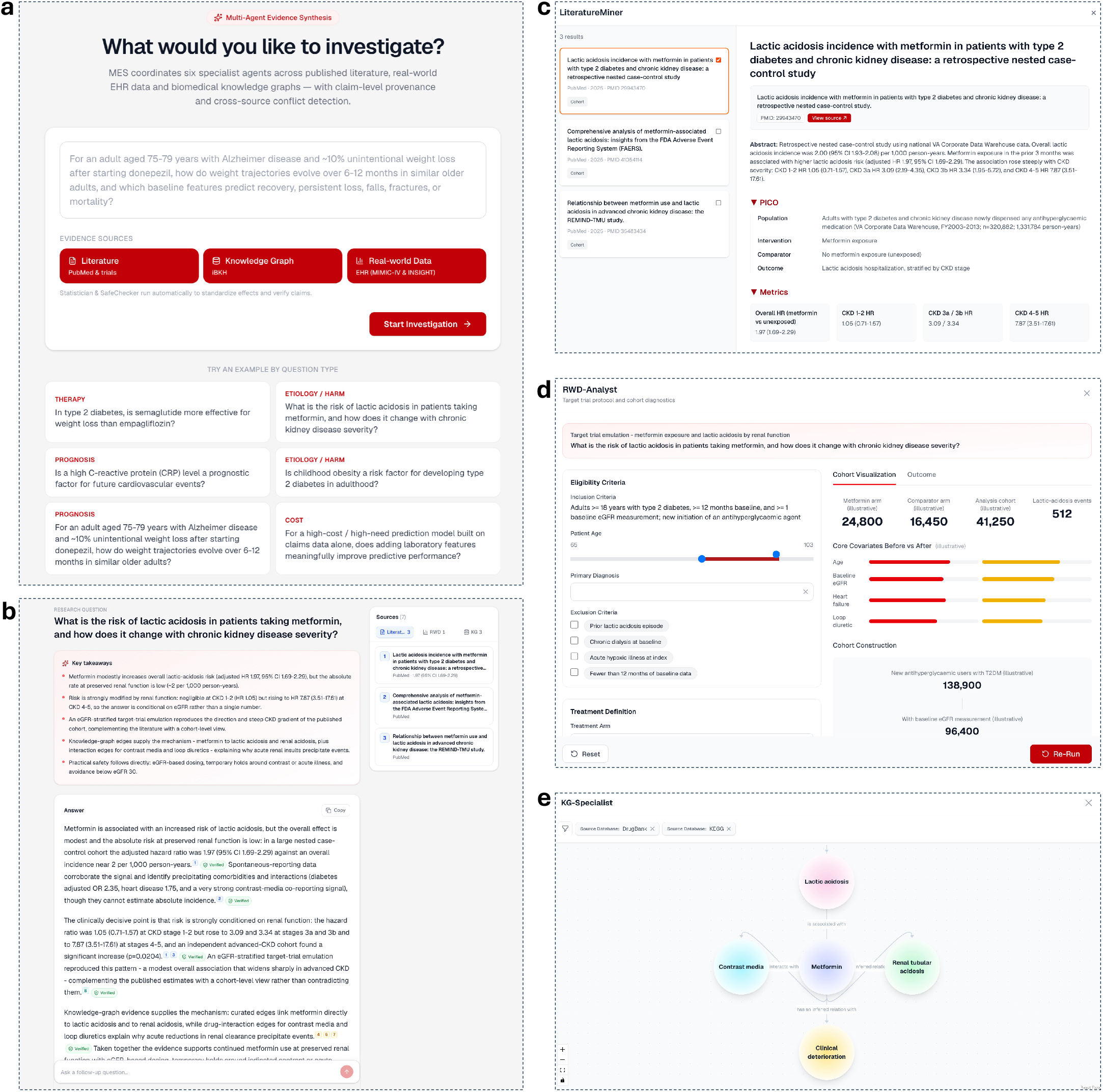
User interface of MES. (a) Query initiation interface. Users enter a natural-language clinical or research question and select evidence sources to be activated, including published literature, knowledge graphs, and real-world clinical data. The interface also provides task-oriented templates for common evidence-based medicine questions, including therapy, etiology/harm, prognosis, and cost-effectiveness. (b) Investigation results interface. After query submission, MES presents a structured investigation summary containing the original research question, source-specific evidence, key findings, and the final synthesized answer with traceable evidence provenance. (c) LiteratureMiner workspace. The interface displays retrieved publications, study details, extracted PICO elements, and quantitative effect estimates, including subgroup-specific metrics when available. (d) RWD-Analyst workspace. The interface supports target trial specification, eligibility criteria, treatment definition, cohort construction, outcome summaries, and covariate balance assessment based on real-world clinical data. (e) KG-Specialist workspace. The interface visualizes biomedical entities and their relationships as an interactive graph, allowing users to inspect source-grounded connections among drugs, diseases, and related clinical concepts. Together, these interfaces expose source-specific evidence and intermediate analytical outputs, making the evidence synthesis process transparent and traceable.

### Results Use Cases

#### Case 1: MES generated real-world evidence when matching literature was absent

We first evaluated MES in a patient-specific clinical management question for which matching published evidence was absent (Fig. 3). The query concerned an adult aged 75–79 years with Alzheimer disease who had developed approximately 10% unintentional weight loss in the six months after initiating donepezil, after common competing causes had been excluded. The clinical question was whether, among clinically similar older adults who developed persistent weight loss after cholinesterase-inhibitor initiation, subsequent weight and safety outcomes differed between those who continued versus discontinued therapy and whether the available evidence could inform switching to an alternative agent.

**Figure 3.**
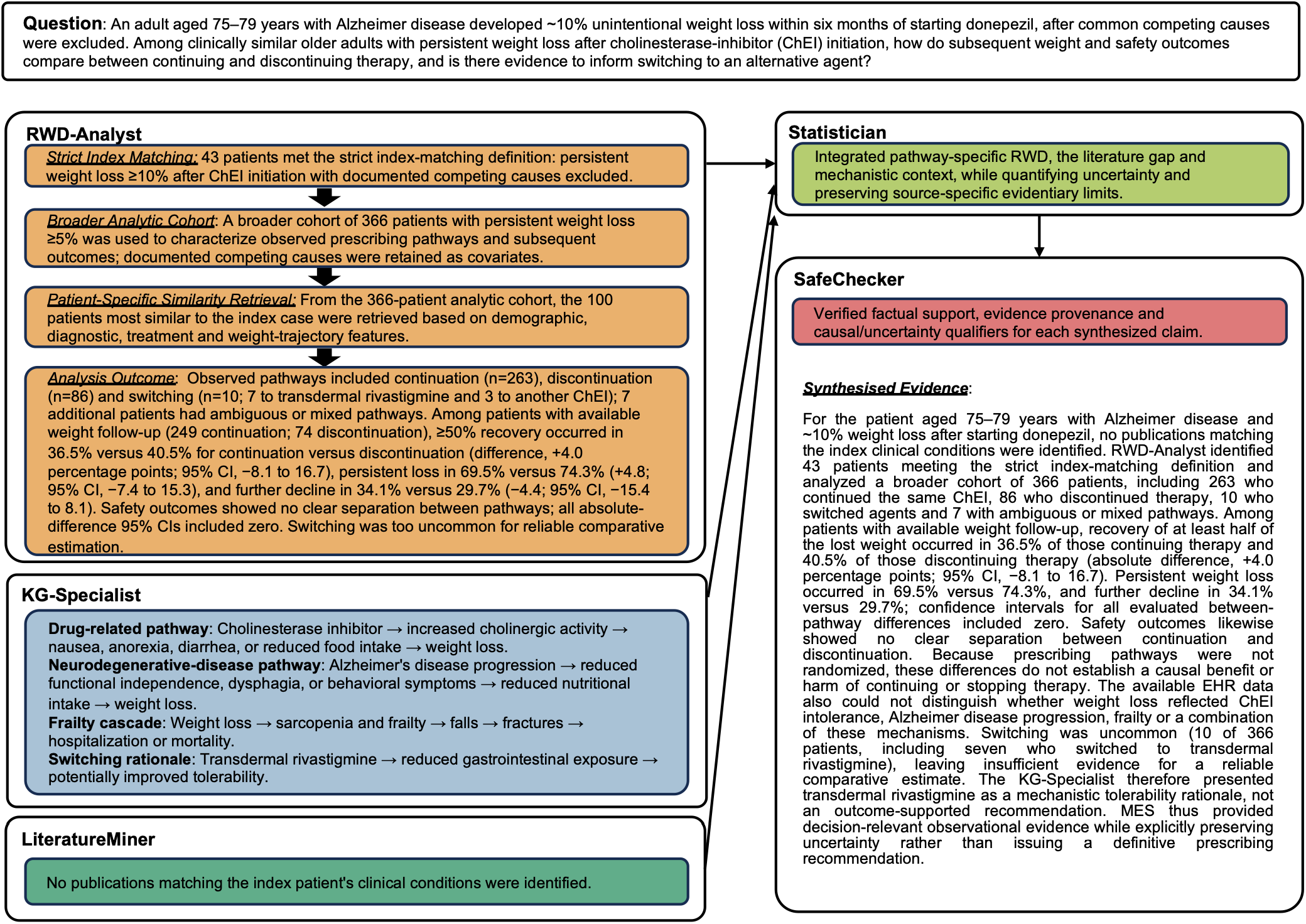
Patient-specific Alzheimer disease management use case. The query asked how clinically similar older adults with persistent weight loss after cholinesterase-inhibitor initiation fared under observed continuation, discontinuation and switching pathways. LiteratureMiner found no matching publications. RWD-Analyst identified 43 patients meeting the strict index-matching definition and analyzed a broader cohort of 366 patients with persistent weight loss of at least 5%; 100 patients most similar to the index case were additionally retrieved for patient-specific context. Pathway-stratified analyses compared weight and safety outcomes between continuation and discontinuation, while switching was reported descriptively because of the small sample. KG-Specialist provided mechanistic context linking cholinesterase-inhibitor intolerance, neurodegenerative disease progression and frailty-related downstream risks. Statistician integrated the evidence and SafeChecker verified claim support.

To illustrate how MES operates when published evidence is sparse, we present a patient-specific clinical management use case (Figure 3). An adult aged 75–79 years with Alzheimer’s disease developed approximately 10% unintentional weight loss within six months of initiating donepezil, with common competing causes excluded. The question was whether outcomes among clinically similar older adults differed between observed continuation and discontinuation pathways and whether the available evidence could inform switching to an alternative agent. LiteratureMiner retrieved no publications matching the index conditions, so the synthesis could not rely on trial- or literature-derived effect estimates. Rather than returning an empty or speculative answer, MES relied on the RWD-Analyst to generate question-specific empirical evidence from real-world data. The RWD-Analyst identified 43 patients meeting the strict index-matching definition and used a broader analytic cohort of 366 patients with persistent weight loss of at least 5% to characterize observed prescribing pathways and outcomes; a similarity-retrieval module additionally identified the 100 patients most similar to the index case. The KG-Specialist supplied mechanistic context linking cholinesterase-inhibitor intolerance, neurodegenerative disease progression and a weight-loss–driven frailty cascade, while the Statistician integrated these heterogeneous outputs and the SafeChecker verified that each claim in the final report was supported by and traceable to its source evidence.

In the broader analytic cohort, 263 patients continued the same cholinesterase inhibitor and 86 discontinued therapy. Among patients with available follow-up weight measurements, recovery of at least half of the lost weight occurred in 91 of 249 patients who continued therapy (36.5%, 95% CI 30.8–42.7%) and 30 of 74 who discontinued therapy (40.5%, 95% CI 30.1–51.9%), corresponding to an absolute difference of 4.0 percentage points (95% CI −8.1 to 16.7). Persistent weight loss occurred in 69.5% versus 74.3% (absolute difference, 4.8 percentage points; 95% CI −7.4 to 15.3), and further decline occurred in 34.1% versus 29.7% (absolute difference, −4.4 percentage points; 95% CI −15.4 to 8.1), respectively. Falls occurred in 16.3% versus 11.6%, fractures in 6.1% versus 3.5%, syncope in 8.4% versus 11.6% and death in 3.8% versus 4.7% in the continuation and discontinuation groups, respectively. Confidence intervals for the absolute between-pathway differences included zero across all evaluated weight and safety outcomes.

This use case highlights several design features of MES that distinguish it from literature-only synthesis tools. First, it demonstrates query-adaptive evidence integration: when one source is uninformative, the system shifts emphasis toward available evidence while reporting the literature gap itself rather than masking it. Second, it preserves the evidentiary boundary of observational comparisons. Because continuation and discontinuation were not randomized, the observed between-pathway differences do not establish a causal effect of continuing or stopping therapy. Moreover, the available EHR data could not determine whether the initial or subsequent weight loss was attributable to cholinesterase-inhibitor intolerance, progression of neurodegenerative disease, frailty or a combination of these mechanisms. Third, switching was uncommon in the broader analytic cohort: 10 of 366 patients switched cholinesterase inhibitors, including seven who switched to transdermal rivastigmine, and no switching was observed among the 100 most similar retrieved patients. The switching sample was therefore insufficient for a reliable comparative estimate, and MES presented the knowledge-graph rationale for transdermal rivastigmine as mechanistic context rather than an outcome-supported recommendation. The resulting report provides observed outcome estimates relevant to continuation versus discontinuation while explicitly identifying the uncertainty surrounding causal attribution and alternative treatment pathways, rather than issuing a definitive prescribing recommendation.

#### Case 2: MES supplied real-world evidence when the literature was inconsistent and inconclusive

We next evaluated MES on a therapeutic adoption question for which published evidence existed but failed to converge (Figure 4). The query concerned whether to adopt short-acting intravenous beta-blockers as an adjunctive treatment for adults with septic shock and persistent tachycardia (heart rate above 95 bpm). It asked what the evidence shows about their effect on patient-important outcomes, principally 28-day mortality. This case is notable because, unlike Case 1, the LiteratureMiner did retrieve directly relevant trials. However, their results conflicted. An early single-center trial suggested a mortality benefit, but this rested on a secondary outcome, an unusually high control-group mortality, and a small open-label design.^48^ The larger multicenter trials did not reproduce it. STRESS-L was stopped early for a signal of possible harm,^49^ and Landi-SEP found no difference in mortality.^50^ The synthesis therefore could not rest on a coherent literature-derived effect estimate. Current guidelines^51^ reflect this deadlock by grading the evidence as very low certainty and flagging beta-blockers in septic shock as an open research priority.

**Figure 4.**
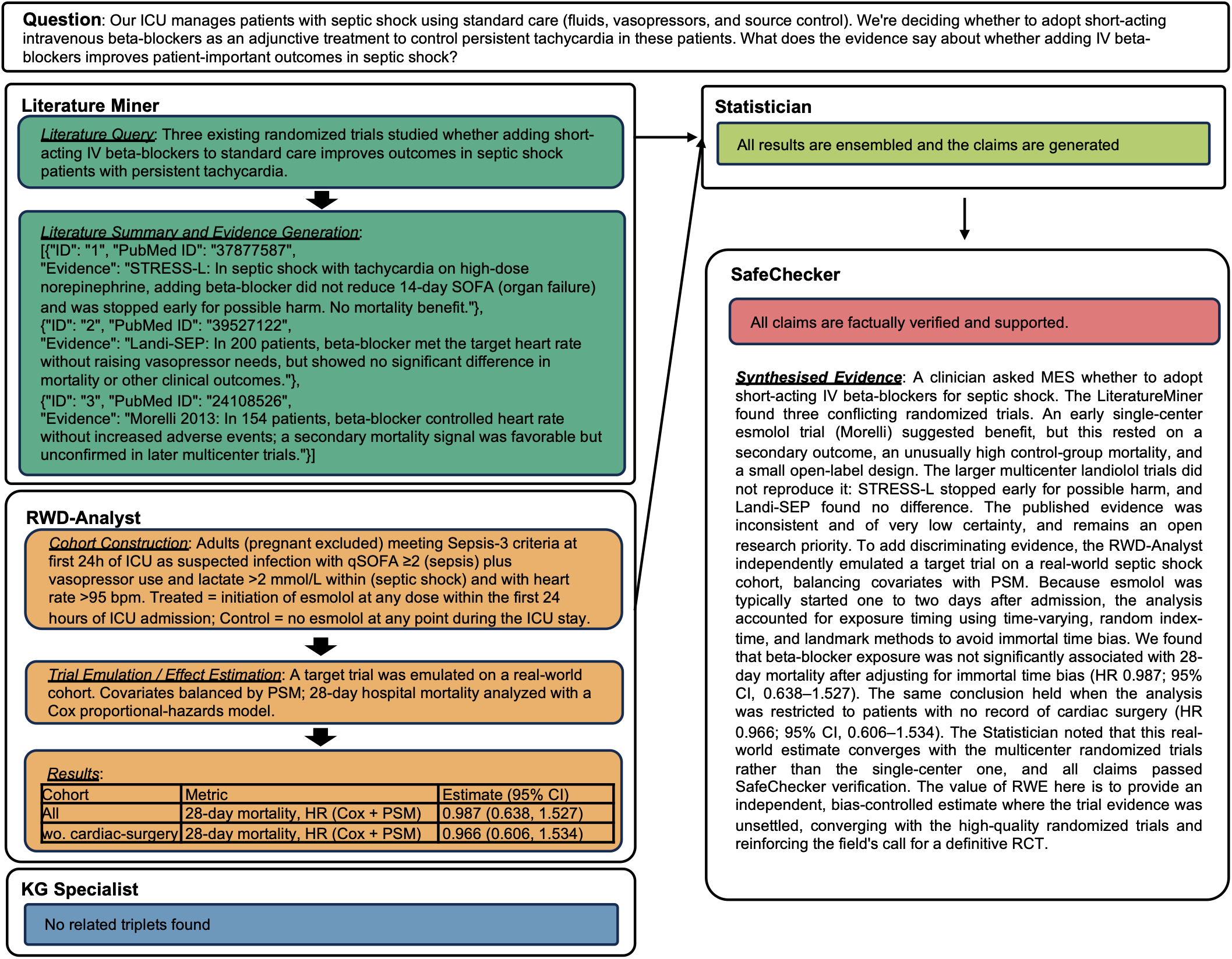
Septic shock beta-blocker adoption use case. The query asked whether adding short-acting intravenous beta-blockers improves outcomes in adults with septic shock and persistent tachycardia (heart rate above 95 bpm), principally 28-day mortality. LiteratureMiner retrieved three relevant randomized trials with conflicting results: an early single-center esmolol trial suggested benefit, while the multicenter landiolol trials showed possible harm (STRESS-L, stopped early) and no difference (Landi-SEP). RWD-Analyst emulated a target trial on a real-world cohort, balancing covariates by PSM and estimating 28-day mortality with a Cox model. Statistician integrated the divergent literature and real-world estimate, and SafeChecker verified claim support.

Rather than propagating the ambiguity or defaulting to the single positive trial, MES used the RWD-Analyst to supply an independent estimate on the contested question. The RWD-Analyst emulated a target trial on a real-world cohort of septic shock patients defining an esmolol-treated group against an untreated control. The emulation was specified transparently. The cohort comprised adults, with pregnant patients excluded, who met Sepsis-3 criteria^52^ within the first 24 hours of ICU admission. These criteria were suspected or documented source of infection combined with an acute increase of 2 points or more in the Sequential Organ Failure Assessment (SOFA) score, vasopressor requirement, and lactate > 2 mmol/L. Patients were further restricted to those with persistent tachycardia, defined as heart rate > 95 bpm, matching the population enrolled in the trials. Treatment was defined as initiation of esmolol at any dose during the first 24 ICU hours. Controls were patients with no esmolol exposure at any point during the ICU stay. A central methodological challenge was that esmolol was typically initiated one to two days after ICU admission, so patients had to survive long enough to receive it. Treating exposure as fixed at baseline would therefore introduce immortal time bias and manufacture an artifactual survival advantage. The RWD-Analyst addressed this directly, estimating the treatment effect after adjusting for immortal time bias by clone-censor-weight methodology. Beta blocker showed no association with 28-day mortality (hazard ratio near 1.0, non-significant). The analysis was then repeated with all patients who had any record of cardiac surgery excluded. This matters because beta-blocker is routine after cardiotomy, so a cohort assembled by septic shock criteria may include patients given esmolol for an entirely different indication. The estimate was essentially unchanged, indicating that the null result was not an artifact of mixed indication. The Statistician integrated the divergent literature with the bias-controlled real-world estimate, and the SafeChecker verified that each claim in the final report was traceable to its source evidence.

This use case highlights a capability that complements Case 1. Case 1 showed MES generating real-world evidence when matching literature was absent. Case 2 shows MES generating an independent real-world estimate when the literature is present but inconsistent. In this setting, the system treats an unresolved trial base not as a dead end but as a signal to seek additional evidence from real-world data. Critically, MES preserves epistemic honesty about how that evidence must be analyzed. It does not simply return a real-world effect estimate. The resulting estimate converges with the multicenter randomized trials rather than the single-center one, reinforcing the guideline’s cautious position and the field’s call for a definitive multicenter trial. This behavior again reflects the system’s intended role as a transparent, source-grounded decision-support framework that complements clinical judgment. Here it does so by adding methodologically rigorous real-world evidence to a question that existing trials have left unsettled, and by guarding against the biases to which observational analyses are most prone.

### Evidence-synthesis quality varied across LLM backbones

We evaluated MES across 144 clinical queries spanning six evidence-based medicine (EBM) categories: clinical effectiveness, comparative effectiveness, diagnostic accuracy, prognosis or risk, safety and adverse events and cost-effectiveness. For each query, MES generated a structured report through coordinated literature retrieval, real-world data analysis, knowledge-graph reasoning, statistical synthesis and claim verification. We evaluated four LLM backbones, GPT-5.4-nano, Gemma 4, MedGemma and Gemma 3, using automated metrics and manual expert assessment.

The four backbones produced distinct synthesis profiles (Fig. 5a–c). MedGemma generated the largest number of evidence items per report and the longest responses, whereas GPT-5.4-nano produced the most concise outputs. All four models achieved high EBM alignment, with mean EBM fractions close to 1.0. The models also differed in their use of qualifying language. GPT-5.4-nano had the lowest hedging ratio, Gemma 4 showed an intermediate profile and MedGemma and Gemma 3 used hedging expressions more frequently.

**Figure 5.**
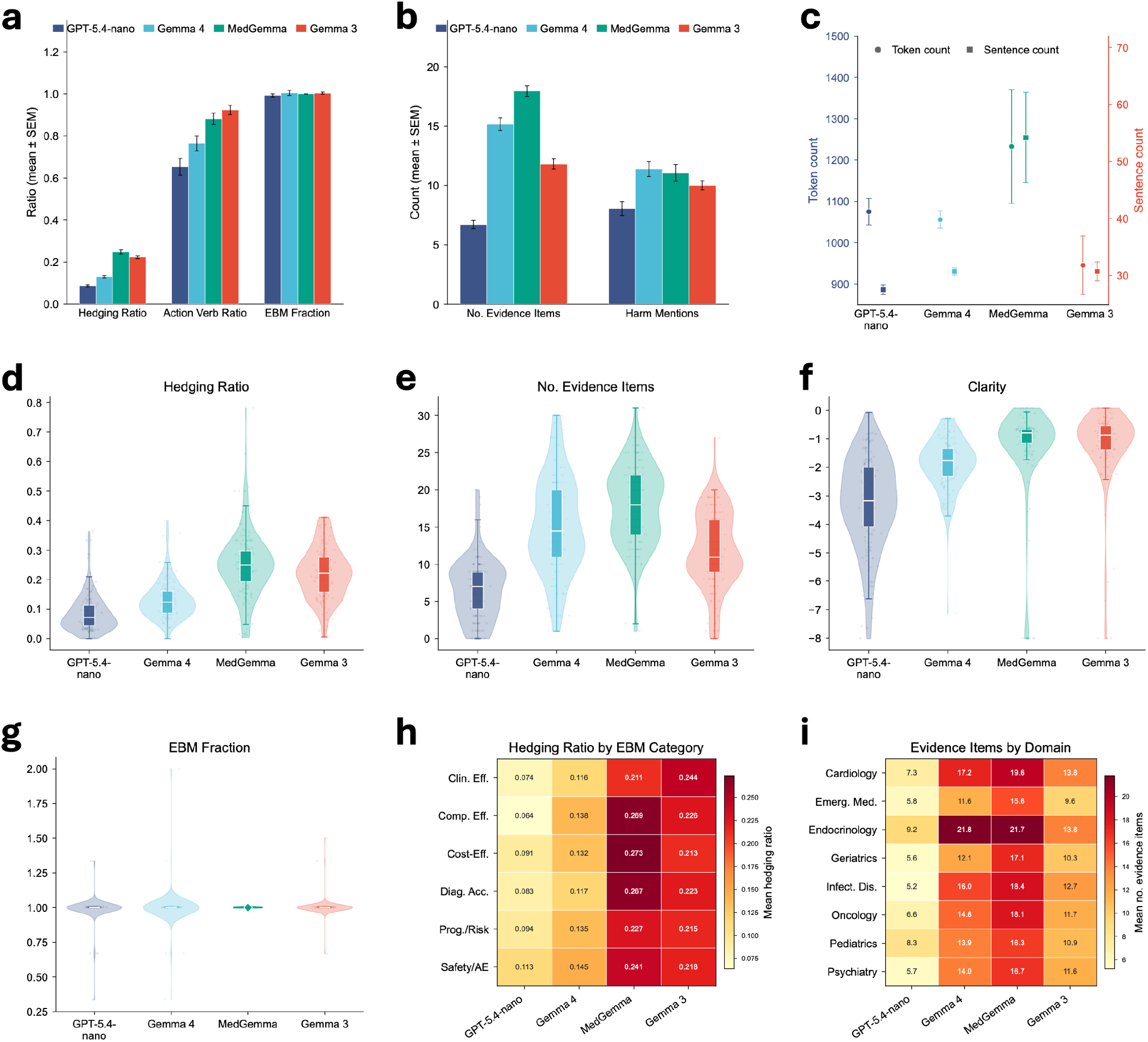
Structural, linguistic and evidence-use evaluation across clinical query types. (a) Mean ratio-based metrics across four LLM backbones, including hedging ratio, action-verb ratio, and evidence-based medicine (EBM) fraction. Error bars indicate SEM across clinical queries. (b) Mean count-based metrics, including the number of evidence items and harm mentions per response. Error bars indicate SEM. (c) Mean response length measured by token count and sentence count, shown on separate y-axes. (d–g) Violin plots showing sample-level distributions of hedging ratio, number of evidence items, clarity proxy, and EBM fraction. Each violin shows the kernel density distribution, embedded box plots indicate the median and interquartile range, and jittered points indicate individual queries. (h) Heatmap of mean hedging ratio stratified by EBM category and LLM backbone. Darker colors indicate more frequent use of uncertainty or qualifying language. (i) Heatmap of mean evidence-item counts stratified by clinical domain and LLM backbone. Darker colors indicate more extensive evidence grounding. Together, these analyses characterize model-specific differences in evidence density, uncertainty communication, safety awareness, readability, and EBM alignment.

Metric definitions are summarized in Table 1. Manual expert assessment showed a different ranking from output length or evidence-item count (Table 2). Across three expert raters, GPT-5.4-nano achieved the highest overall score (4.51 ± 0.48), followed by Gemma 4 (4.00 ± 0.35), Gemma 3 (2.79 ± 0.57) and MedGemma (2.75 ± 0.61). Differences across models were significant for every evaluated dimension and for the overall score (Kruskal-Wallis test, all P < 0.001). GPT-5.4-nano received the highest ratings for addressing the clinical question, identifying clinical elements, using appropriate evidence, distinguishing evidence strength, explaining the evidence-to-conclusion link, acknowledging uncertainty, avoiding overclaiming, providing actionable interpretation and communicating safety considerations.

**Table 1.** Definitions and interpretation of structural, linguistic, and evidence-use metrics for evidence synthesis evaluation. The table defines the metrics used to characterize model-generated evidence synthesis reports across four LLM backbones. For each metric, we describe what it measures, how it was calculated, and how higher or lower values should be interpreted. The metrics cover uncertainty communication, actionability, alignment with evidence-based medicine (EBM) categories, evidence grounding, safety awareness, response length, readability, and subgroup-level variation across EBM categories and clinical domains. These metrics were used to complement manual expert ratings by quantifying differences in evidence density, cautious language, clinical risk communication, readability, and domain-specific evidence coverage.

| Metric | What it measures | Calculation | Interpretation |
| --- | --- | --- | --- |
| **Hedging ratio** | Degree of uncertainty or cautious language in the response | Number of hedging expressions divided by total sentence or token units, depending on implementation. Hedging terms include expressions such as “may,” “might,” “could,” “suggests,” “appears,” “limited evidence,” and “uncertain.” | Higher values indicate more cautious or qualified language. This can be appropriate when evidence is heterogeneous, but excessive hedging may reduce decisiveness. |
| **Action-verb ratio** | Density of clinically actionable language | Number of action-oriented verbs or phrases divided by total sentence or token units. Examples include “monitor,” “assess,” “evaluate,” “consider,” “compare,” “screen,” and “adjust.” | Higher values indicate that the response provides more actionable recommendations or next steps. |
| **EBM fraction** | Alignment with predefined evidence-based medicine categories | Ratio of identified EBM-aligned clinical elements to the expected elements defined by the applicable EBM framework, including clinical effectiveness, comparative effectiveness, diagnostic accuracy, prognosis/risk, safety/adverse events and cost-effectiveness. | Values close to 1 indicate that the expected EBM elements are represented. Values below 1 indicate lower coverage, whereas values slightly above 1 can occur when additional or repeated relevant clinical elements are identified. |
| **Number of evidence items** | Breadth of cited or summarized evidence | Count of distinct evidence items extracted, cited, or summarized in each response. Evidence items may include studies, trials, real-world-data analyses, knowledge-graph relationships, or structured evidence statements. | Higher values indicate richer evidence grounding, although very high counts may reflect evidence piling rather than concise synthesis. |
| **Harm mentions** | Safety-awareness of the response | Count of words or phrases related to harms, adverse events, risks, toxicity, contraindications, side effects, complications, or safety limitations. | Higher values indicate greater attention to clinical risks and safety considerations, especially important for treatment and safety-related questions. |
| **Token count** | Overall response length | Total number of tokens in each generated response. | Higher values indicate longer responses. This may reflect more comprehensive synthesis but can also reduce readability. |
| **Sentence count** | Structural length of the response | Total number of sentences in each generated response. | Higher values indicate more segmented explanations. Combined with token count, this helps distinguish concise multi-sentence reports from dense long-sentence reports. |
| **Clarity** | Readability of the response | A Flesch-style readability metric computed from sentence length and word complexity, clipped to the 2nd–98th percentile for visualization. In this implementation, more negative values indicate lower readability, while values closer to zero indicate clearer text. | Higher or less negative values indicate easier-to-read outputs. More negative values suggest denser, more technical, or harder-to-interpret language. |
| **Mean hedging ratio by EBM category** | Whether uncertainty varies across clinical question types | Average hedging ratio computed within each EBM category and model backbone. | Identifies categories where models are more cautious. Prognostic/risk and safety/adverse-event questions often show higher hedging because evidence is more heterogeneous. |
| **Mean evidence items by clinical domain** | Whether evidence grounding varies across medical specialties | Average number of evidence items computed within each clinical domain and model backbone. | Identifies domains where responses are more evidence-dense or more sparse, revealing possible domain-specific strengths and weaknesses. |

**Table 2.** Manual expert assessment of evidence synthesis reports generated with four LLM backbones. Three expert raters evaluated reports across 21 criteria grouped into seven dimensions: clinical relevance, evidence appropriateness, reasoning quality, uncertainty handling, usefulness and interpretability, actionability and safety awareness. Scores are reported as mean ± standard deviation. P-values were calculated using the Kruskal–Wallis test^53^. The test was non-directional, and no adjustment for multiple comparisons was applied. GPT-5.4-nano achieved the highest average score across all criteria, with statistically significant differences across models for every individual criterion and for the overall average score.

| Category | Criterion | GPT-5.4 nano | Gemma 4 | MedGemma | Gemma 3 | P-Value | Best Model |
| --- | --- | --- | --- | --- | --- | --- | --- |
| Clinical Relevance | It directly addresses the clinical question. Score higher if the response clearly answers the specific clinical question. Score lower if it is vague, off-topic, or only partially related. | 4.65±0.46 | 4.18±0.42 | 3.59±0.64 | 3.50±0.84 | < 0.001 | GPT-5.4 nano |
|  | It identifies key clinical elements. Score higher if the response identifies key elements such as population, intervention/exposure, comparator, outcome, or clinical setting when applicable. | 4.69±0.43 | 4.26±0.34 | 3.65±0.60 | 3.53±0.84 | < 0.001 | GPT-5.4 nano |
|  | It provides clinically meaningful conclusions. Score higher if the response gives specific and clinically meaningful conclusions rather than generic statements. | 4.58±0.52 | 4.09±0.38 | 3.45±0.58 | 3.20±0.75 | < 0.001 | GPT-5.4 nano |
| Evidence Appropriateness | It uses relevant evidence. Score higher if the evidence cited or discussed is directly relevant to the clinical question. | 4.54±0.70 | 4.12±0.42 | 2.74±0.53 | 2.86±0.60 | < 0.001 | GPT-5.4 nano |
|  | It distinguishes evidence strength. Score higher if the response distinguishes stronger evidence from weaker, indirect, observational, or mechanistic evidence. | 4.57±0.56 | 4.04±0.34 | 2.66±0.47 | 2.78±0.58 | < 0.001 | GPT-5.4 nano |
|  | It considers evidence applicability. Score higher if the response considers whether the evidence applies to the target population, setting, intervention, or outcome. | 4.45±0.47 | 4.00±0.29 | 2.61±0.43 | 2.55±0.47 | < 0.001 | GPT-5.4 nano |
| Reasoning Quality | It explains evidence-to-conclusion reasoning. Score higher if the response clearly explains how the evidence supports the final conclusion. | 4.73±0.47 | 3.96±0.39 | 2.56±0.48 | 2.55±0.53 | < 0.001 | GPT-5.4 nano |
|  | It interprets quantitative or comparative evidence. Score higher if the response correctly interprets effect estimates, confidence intervals, statistical significance, or comparative findings. | 4.66±0.58 | 4.05±0.38 | 2.63±0.46 | 2.55±0.49 | < 0.001 | GPT-5.4 nano |
|  | It considers bias, confounding, and heterogeneity. Score higher if the response considers bias, confounding, study limitations, heterogeneity, or population differences when relevant. | 4.37±0.41 | 3.78±0.26 | 2.58±0.42 | 2.52±0.43 | < 0.001 | GPT-5.4 nano |
| Uncertainty Handling | It acknowledges limitations and uncertainty. Score higher if the response clearly states important limitations, uncertainty, missing evidence, or evidence gaps. | 4.64±0.29 | 3.85±0.32 | 2.79±0.42 | 2.72±0.37 | < 0.001 | GPT-5.4 nano |
|  | It identifies conflicting evidence. Score higher if the response recognizes disagreement across studies, populations, methods, or evidence sources when present. | 4.23±0.30 | 3.68±0.21 | 2.63±0.41 | 2.57±0.37 | < 0.001 | GPT-5.4 nano |
|  | It avoids overclaiming. Score higher if the response uses cautious conclusions when evidence is limited, mixed, indirect, or low quality. | 4.76±0.28 | 3.88±0.31 | 2.77±0.44 | 2.71±0.38 | < 0.001 | GPT-5.4 nano |
| Usefulness and Interpretability | It is useful for clinical or research decision-making. Score higher if the response helps clinicians, researchers, or decision-makers understand and use the evidence. | 4.69±0.43 | 4.26±0.32 | 2.72±0.46 | 2.73±0.52 | < 0.001 | GPT-5.4 nano |
|  | It provides implications, cautions, or next steps. Score higher if the response provides clear implications, cautions, or suggestions for further investigation when appropriate. | 4.63±0.40 | 4.05±0.26 | 2.66±0.46 | 2.63±0.49 | < 0.001 | GPT-5.4 nano |
|  | It is well-structured and easy to interpret. Score higher if the response is logically organized, readable, and easy to follow. | 4.65±0.35 | 4.21±0.22 | 2.73±0.40 | 2.67±0.45 | < 0.001 | GPT-5.4 nano |
| Actionability | It provides actionable interpretation. Score higher if the response helps users understand what clinical, research, or decision-making actions could be considered. | 4.59±0.55 | 4.08±0.35 | 2.67±0.46 | 2.64±0.50 | < 0.001 | GPT-5.4 nano |
|  | It clarifies when evidence is insufficient for action. Score higher if the response clearly states when evidence is not strong enough to support clinical action or research conclusions. | 4.36±0.28 | 3.78±0.25 | 2.57±0.41 | 2.52±0.47 | < 0.001 | GPT-5.4 nano |
|  | It supports future evidence generation. Score higher if the response helps guide study design, cohort definition, outcome selection, subgroup analysis, or future investigation. | 4.38±0.36 | 3.90±0.17 | 2.66±0.46 | 2.64±0.50 | < 0.001 | GPT-5.4 nano |
| Safety Awareness | It identifies safety concerns. Score higher if the response identifies relevant adverse events, contraindications, safety signals, or potential harms when applicable. | 4.01±0.41 | 3.93±0.33 | 2.66±0.45 | 2.58±0.49 | < 0.001 | GPT-5.4 nano |
|  | It communicates risk carefully. Score higher if the response communicates safety risks clearly, proportionally, and without exaggeration or minimization. | 4.23±0.34 | 3.94±0.30 | 2.64±0.41 | 2.60±0.46 | < 0.001 | GPT-5.4 nano |
|  | It distinguishes efficacy from safety. Score higher if the response separates evidence about benefit or effectiveness from evidence about safety or harm. | 4.23±0.34 | 3.88±0.27 | 2.64±0.42 | 2.59±0.46 | < 0.001 | GPT-5.4 nano |
| Average Score |  | 4.51±0.48 | 4.00±0.35 | 2.79±0.57 | 2.75±0.61 | < 0.001 | GPT-5.4 nano |

These findings indicate that synthesis quality depends on evidence selection, compression and communication, not only on the amount of evidence retrieved. Although MedGemma produced the most evidence-dense reports, GPT-5.4-nano generated shorter reports with higher expert ratings for clinical relevance, reasoning quality, actionability and safety-aware communication.

### Cross-Source Evidence Integration and Evidence Coverage Across LLM Backbones

We next evaluated cross-source integration using automated metrics for evidence volume, integration efficiency, EBM alignment and safety-related language (Fig. 6). MES synthesizes multi-source evidence by processing original evidence items into normalized summaries. The normalization ratio was calculated as the number of normalized evidence items divided by the number of original evidence items. Lower values indicate greater compression of the original evidence set, whereas higher values indicate greater retention of evidence items after normalization.

**Figure 6.**
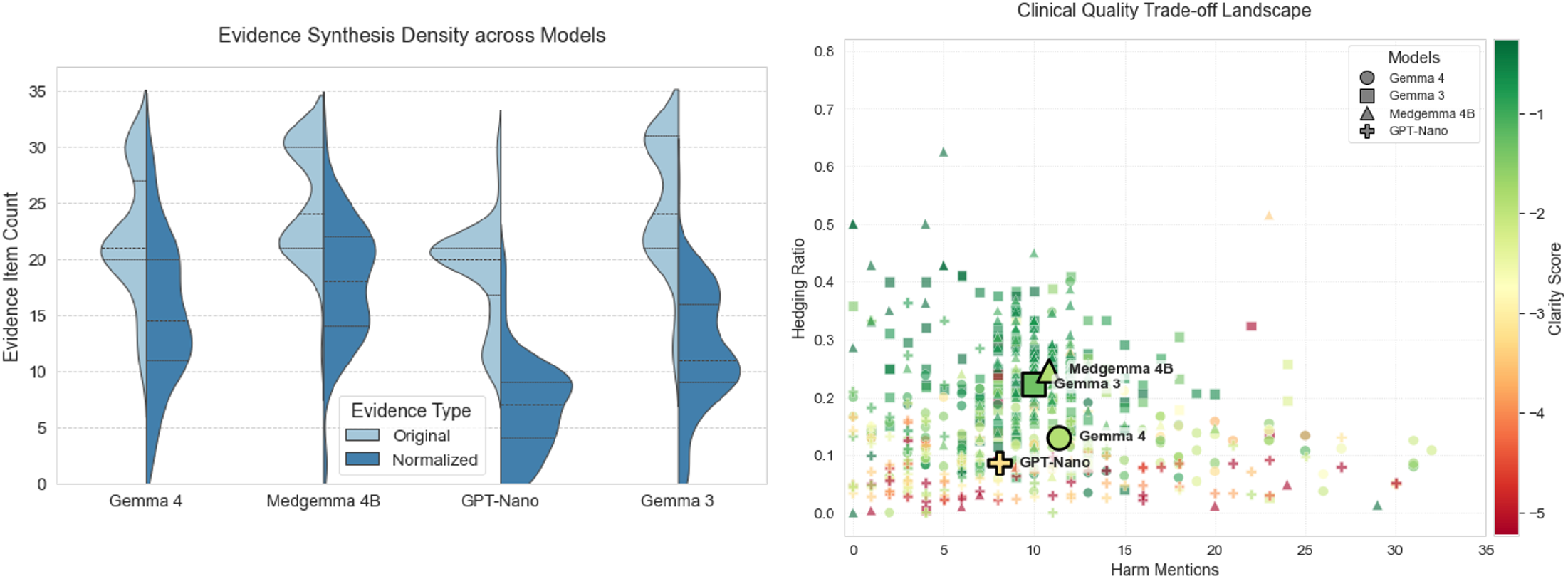
Cross-source evidence integration and clinical-quality trade-off landscape. The left panel compares original and normalized evidence-item counts across model backbones. The right panel relates harm mentions, hedging ratio and clarity score across generated reports, showing how evidence density, safety-related language, uncertainty communication and readability vary across backbones.

In our evaluation, Gemma 4 processed an average of 22.10 original evidence items per query into 15.16 normalized evidence summaries, corresponding to a normalization ratio of 68.6%. In comparison, GPT-5.4-nano processed 18.92 original items into 6.70 normalized items, corresponding to a normalization ratio of 35.4%. These results indicate different synthesis profiles across backbones, with GPT-5.4-nano applying greater compression and Gemma 4 retaining a larger proportion of the original evidence items after normalization. MedGemma produced the highest volume of normalized evidence (17.94 items). Alignment with the EBM framework is summarized by the EBM fraction. Values near 1.00 indicate that the expected clinical elements are represented in the response. Values slightly above 1 can occur when responses contain additional or repeated instances of relevant clinical elements, such as multiple interventions or outcomes. Gemma 4 achieved an average EBM fraction of 1.0046, closely followed by Gemma 3 (1.0031), MedGemma (1.00) and GPT-5.4-nano (0.9931). Notably, all four models successfully maintained a 100% internal consistency rate across their multi-agent outputs. A critical component of clinical evidence synthesis is its focus on safety and risk communication. The frequency of “harm-mentions” (e.g., toxicity, side effects, contraindications) serves as a proxy for how well a framework prioritizes safety-critical information. Analysis of this metric showed Gemma 4 averaging 11.38 terms per summary, followed by MedGemma (10.82), Gemma 3 (9.99), and GPT-5.4-nano (8.06).

Together, these findings characterize how MES processes heterogeneous evidence across LLM backbones. Literature, RWD and knowledge graphs contribute distinct forms of evidence, while the observed normalization, evidence-use and consistency metrics describe how these sources are represented within the resulting multi-source reports.

### MES Detected and Characterized Evidence Divergence

The ability to detect and flag divergence between diverse evidence sources, such as contradictions between clinical trials and real-world data, is a primary safety mechanism in automated synthesis. A higher divergence detection rate indicates that a model successfully identifies discrepancies rather than forcing an artificial consensus. In our evaluation, Gemma 3 and Gemma 4 demonstrated high sensitivity to these discrepancies, identifying contradictions in 86.11% and 83.33% of queries, respectively. In contrast, GPT-5.4-nano detected divergences in 68.06% of queries, and MedGemma identified them in 54.17%. These metrics underscore the functional impact of a dedicated verification module like SafeChecker for parsing nuanced evidence divergences.

Once a divergence was identified, models differed in how they represented it in the final synthesis. Retaining the disagreement rather than collapsing it into a single conclusion preserves transparency about unresolved evidence. Gemma 4 retained the detected divergence in 115 of 120 flagged queries (95.8%). MedGemma retained 70 of 78 detected divergences (89.7%) and synthesized the remaining 8 cases (10.3%). Overall, the models more often retained cross-source disagreement than collapsed it into a single consensus.

The communication of these findings is evaluated using the hedging ratio, which measures the directness of the generated text. A lower hedging ratio indicates clear, decisive language, whereas a higher ratio points to a reliance on qualifying or non-committal phrasing. MES with Gemma 4 produced a hedging ratio of 0.131, generating more direct summaries than MedGemma (0.247) and Gemma 3 (0.223). This demonstrates an ability to be decisive when evidence aligns, while relying on explicit conflict reporting when it does not. However, directness must always be balanced with interpretability. GPT-5.4-nano exhibited the lowest hedging ratio (0.087) but concurrently scored the lowest in clarity (-3.231), indicating that its outputs were difficult to interpret despite lacking qualifying language.

**Figure 7.**
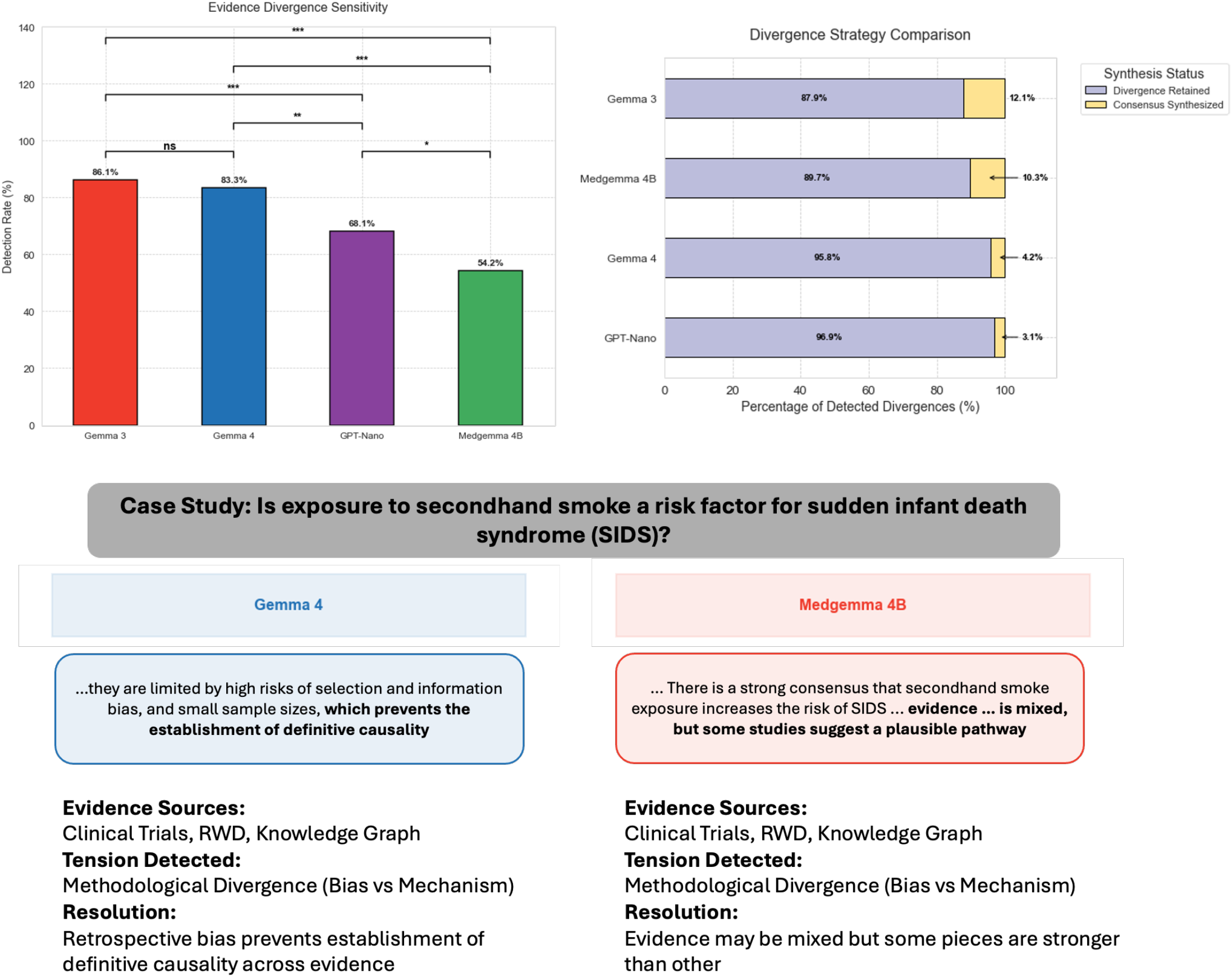
Identification and handling of conflicting evidence. The upper-left panel shows divergence-detection rates across LLM backbones. The upper-right panel shows whether detected divergences were retained in the final synthesis or collapsed into consensus. The lower panel illustrates a case study on secondhand smoke exposure and sudden infant death syndrome, where different backbones used different strategies for communicating conflicting evidence. ns, P ≥ 0.05; *P < 0.05; **P < 0.01; ***P < 0.001.

### SafeChecker Improved Claim-evidence Verification

We evaluated SafeChecker on CliniFact^54^, a benchmark for claim–evidence verification in clinical research. CliniFact contains 1,970 claim–evidence pairs derived from 992 unique interventional clinical trials and linked to 1,540 scientific publications. Each claim is constructed from a trial’s primary outcome, intervention, comparator and statistical hypothesis, and is paired with an abstract from the corresponding publication. Each pair is labelled as Evidence when the abstract supports the claim, Inconclusive when the reported finding is inconclusive, or not enough information (NEI) when the abstract does not contain sufficient relevant information to assess the claim. Because each instance contains one claim and one source abstract, the benchmark directly evaluates whether SafeChecker can determine if an individual claim is grounded in its cited evidence.

We compared two verification conditions on an evaluation set of 394 claim–evidence pairs. The first used direct prompting with the classification prompt from the original CliniFact study (see Supplementary Note 7). The second used SafeChecker as the verification agent. We first evaluated three-class classification across the original CliniFact labels, as shown in Supplementary Fig. 1. We then evaluated binary unsupported-claim detection by grouping Inconclusive and NEI instances as non-supporting evidence. These instances were treated as the positive class, whereas Evidence instances were treated as the negative class. Performance was evaluated using accuracy, precision, recall and F1 score.

SafeChecker outperformed direct prompting across all four binary classification metrics (Supplementary Table 1). Accuracy increased from 0.850 to 0.926, precision from 0.865 to 0.926, recall from 0.936 to 0.975 and F1 score from 0.899 to 0.950. Consistent with these aggregate results, the three-class confusion matrices showed that SafeChecker reduced the total number of classification errors from 74 to 44. The largest improvements were observed for the Inconclusive and Evidence classes, while performance for NEI remained comparable (Supplementary Fig. 1). These results indicate that a dedicated verification agent improves detection of claims that are not supported by their cited evidence. By tracing individual claims to their source abstracts and evaluating whether each source directly supports the stated conclusion, SafeChecker reduces unsupported statements and makes the evidentiary basis of the final synthesis more explicit. This verification step is particularly important in medical evidence synthesis, where unsupported or overstated claims could lead to misleading clinical interpretations.

### Sensitivity Analyses

We examined whether differences across LLM backbones were consistent at the individual-query level and across clinical contexts (Fig. 5d–i). Query-level distributions showed clear differences in report characteristics. GPT-5.4-nano consistently produced the lowest hedging ratios. MedGemma and Gemma 3 used qualifying language more frequently. MedGemma generated the largest median number of evidence items and had a broader upper tail, followed by Gemma 4. GPT-5.4-nano produced fewer evidence items per report. The clarity proxy also varied. GPT-5.4-nano had more negative scores, whereas MedGemma and Gemma 3 had scores closer to zero on average. EBM fractions remained tightly concentrated around 1.0 across all four models.

Hedging ratios also varied by EBM category. GPT-5.4-nano had the lowest hedging ratio across all six categories. MedGemma and Gemma 3 generally showed higher values. Safety and adverse-event questions showed relatively high hedging ratios for GPT-5.4-nano and Gemma 4, whereas MedGemma showed higher ratios for comparative-effectiveness, cost-effectiveness and diagnostic-accuracy questions.

Evidence-item counts varied across clinical domains. MedGemma generated the largest number of evidence items across all eight domains, while GPT-5.4-nano generated the fewest. The largest differences were observed in endocrinology, where mean evidence-item counts ranged from 9.2 for GPT-5.4-nano to 21.7 for MedGemma, and in cardiology, where values ranged from 7.3 to 19.6. Gemma 4 and Gemma 3 generally showed intermediate values. These analyses confirm that backbone-level differences were not limited to aggregate averages.

## Discussion

MES extends automated evidence synthesis beyond literature summarization toward auditable, multi-source clinical reasoning. Its central contribution is not that a language model can produce a longer report. Rather, MES separates evidence retrieval, real-world analysis, knowledge-graph reasoning, statistical integration and verification into explicit roles with traceable outputs. This design preserves distinctions among trial evidence, observational findings and mechanistic context while still producing a single structured synthesis for the user.

The evaluations show that synthesis quality depends on how evidence is selected, compressed and communicated. Across 144 clinical queries, all four LLM backbones maintained high alignment with predefined EBM categories, but they differed substantially in report length, evidence density, hedging and expert-rated quality. GPT-5.4-nano received the highest mean expert score, whereas MedGemma produced the most evidence-dense reports. This contrast indicates that evidence accumulation alone is not sufficient. Retrieved items must be organized around the clinical question, evidence strength, applicability and decision boundary. For medical interpretation, concise reasoning with explicit uncertainty may be more useful than exhaustive summarization.

The use cases illustrate why source-adaptive synthesis matters. In the Alzheimer disease management example, the literature search did not identify matching publications. MES therefore relied on real-world data to compare observed continuation and discontinuation pathways with explicit uncertainty, used knowledge-graph evidence as mechanistic context and reported that the small switching sample was insufficient for a reliable comparative estimate. In the septic shock beta-blocker example, directly relevant randomized trials were available but produced inconsistent findings. MES therefore supplemented the unresolved literature with an independent real-world analysis and preserved the cross-source disagreement in the final synthesis. The real-world estimate was more consistent with the larger multicenter trials than with the earlier single-center study. Together, these examples demonstrate two behaviours that are difficult for single-source summarizers: treating missing evidence as an informative result and using additional evidence to contextualize unresolved disagreement without forcing consensus.

Cross-source divergence handling is a second important function of MES. Automated synthesis can create clinical risk when it smooths over discordant evidence or presents forced consensus. MES instead represents disagreement as an object of analysis. Statistician examines whether divergence may reflect study design, population, measurement, confounding or heterogeneity. SafeChecker then verifies that the final report does not turn unresolved divergence into a stronger conclusion than the evidence supports. In the benchmark, backbones differed in divergence sensitivity, with Gemma 3 and Gemma 4 detecting contradictions more often than GPT-5.4-nano and MedGemma. Most detected divergences were retained rather than collapsed into consensus. This behavior is useful because unresolved conflict is often a clinically meaningful output.

Claim-level verification is essential for clinical applications of LLM-based synthesis. SafeChecker improved performance on CliniFact compared with direct prompting, increasing accuracy from 0.850 to 0.926 and F1 score from 0.899 to 0.950. This result supports a dedicated verification layer that decomposes reports into testable claims and checks each claim against cited evidence. The theoretical analysis frames SafeChecker as a calibrated screening mechanism rather than an oracle. It can bound false-claim acceptance under stated assumptions, but it cannot guarantee that upstream evidence is complete, unbiased or clinically applicable. Verification reduces unsupported statements, but it does not remove the need to assess data quality, source coverage and applicability.

Several limitations define the current boundary of MES. First, the quality of the final synthesis remains constrained by the quality, coverage and accessibility of its underlying evidence sources. EHR-based analyses are susceptible to missing data, measurement error, coding variation, unmeasured confounding^55^ and limited follow-up,^56^ while automated concept mapping and computable phenotype construction were not independently validated for every query and may introduce cohort misclassification. The applicability of generated RWE is also limited by the populations, healthcare systems and time periods represented in the underlying data. In particular, MIMIC-IV is derived from a single U.S. academic medical center, and evidence generated from this resource should not be assumed to generalize to other institutions or geographic settings. The current evaluation also does not include independent external validation of each generated causal or predictive analysis, leaving the transportability of these estimates across datasets to be established. Second, although MES explicitly considers study design, risk of bias, precision, applicability and cross-source disagreement when synthesizing evidence, it does not implement a formal evidence-certainty framework such as GRADE.^57,58^ The resulting synthesis should therefore not be interpreted as providing a standardized certainty rating or recommendation strength. Knowledge graphs may additionally omit relevant relations or encode associations that are not causal.^59,60^ Literature retrieval may miss unpublished evidence, negative studies or studies outside the indexed databases used by the system. The choice of LLM backbone also affects synthesis style and expert-rated quality, indicating that the architecture does not eliminate model dependence.^61^ Finally, the present evaluation assesses report quality, evidence handling and claim verification, but does not establish prospective clinical benefit, improvement in decision-making or equivalence to a full systematic review.

Future work should therefore focus on external, task-specific and workflow-level validation. Important directions include independently validating generated cohorts and computable phenotypes, replicating causal and predictive analyses across institutions and datasets, and evaluating MES in clinical and research workflows to determine whether it improves evidence-review efficiency or decision quality. Incorporating more formal approaches to evidence-certainty assessment, including frameworks such as GRADE^57,58^, could further standardize how risk of bias, inconsistency, indirectness, imprecision and other limitations are communicated across evidence sources. Additional work should expand source coverage beyond the current literature, EHR and knowledge-graph resources, strengthen calibration of claim verification across dependent claims within a report and define principled stopping criteria for evidence-exhausted states. The appropriate role for MES remains that of a transparent evidence-generation and synthesis framework that helps users distinguish what is supported, uncertain, conflicting or absent while leaving final clinical and policy judgement to domain experts.

## Data Availability

The INSIGHT data can be requested through https://insightcrn.org/. The de-identified data utilized in this study MIMIC-IV can be accessed upon the approval of a formal proposal and the execution of a Data Access Agreement via PhysioNet (https://physionet.org/).

## Code Availability

For reproducibility, our codes are available at GitHub https://github.com/TrialLab/MES. All implementation and analysis details are documented within the repository.

## Acknowledgements

We would like to acknowledge the support from NSF 2212175, NIH RF1AG072449, RF1AG084178, R01AG080991, R01AG080624, R01AG076448, R01AG076234, and R01NS140142.

## Author Contributions Statement

F.W. conceived the initial idea. H.L. led the methodological development and computational experiments. H.L., W.P. and S.R. jointly implemented the method and software. H.L. and S.R. performed the overall statistical analyses, and W.P. conducted the additional statistical analysis of SafeChecker. W.P., H.L., J.G., V.L., V.F., M.P., L.L., A.J., and A.B. developed the user interface. H.L., W.P. and S.R. drafted the initial manuscript, with critical revisions by F.W. C.Z., H.Y., and E.S. reviewed the manuscript and provided suggestions. F.W. supervised the project. All authors reviewed, provided feedback, and approved the final manuscript.

