## Supplementary Information for "MES: A Multi-Agent Evidence Synthesis System for Medical Decision-Making"

**Supplementary Table 1. SafeChecker performance on CliniFact claim–evidence verification.** The table compares direct prompting and SafeChecker on the 394-pair CliniFact evaluation set using accuracy, precision, recall and F1 score. For binary unsupported-claim detection, claim–evidence pairs in which the cited abstract did not support the corresponding claim were treated as positive samples. This positive class included both Inconclusive and NEI instances; Evidence instances were treated as the negative class.

| | Accuracy | Precision | Recall | F1 Score |
|---|---|---|---|---|
| Baseline | 0.850 | 0.865 | 0.936 | 0.899 |
| SafeChecker | 0.926 | 0.926 | 0.975 | 0.950 |

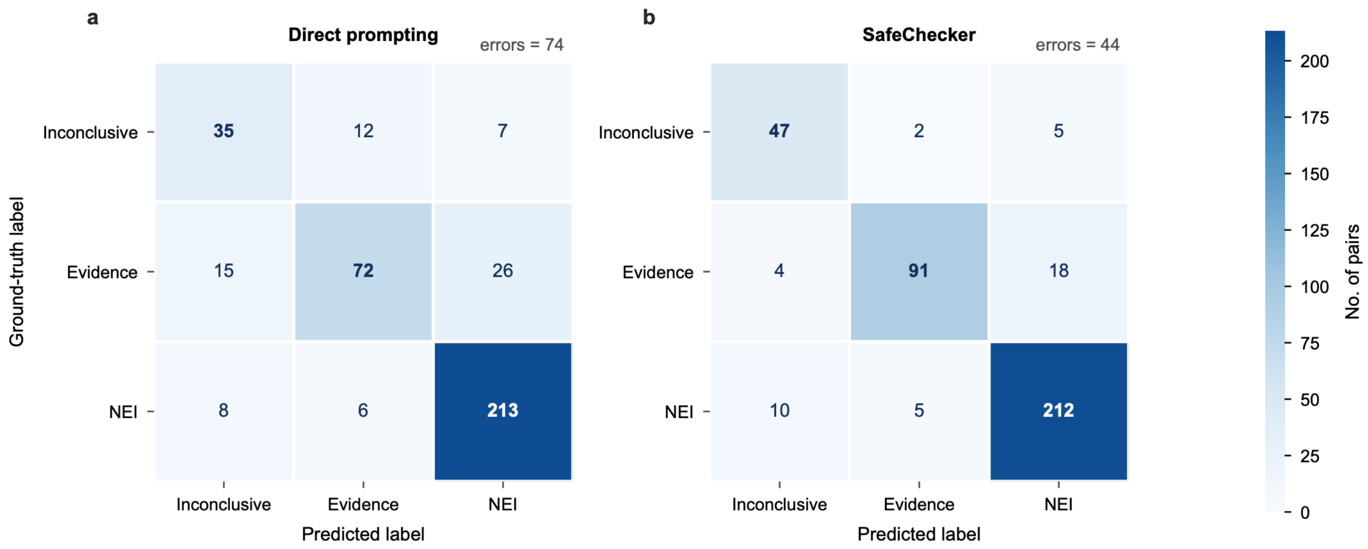


**Supplementary Figure 1. SafeChecker improves claim–evidence verification on the CliniFact benchmark.** Confusion matrices compare predictions obtained using (a) direct prompting with the classification prompt from the original CliniFact study and (b) SafeChecker on the 394-pair CliniFact evaluation set. Rows show ground-truth labels and columns show predicted labels for the three CliniFact classes: Inconclusive, indicating inconclusive evidence; Evidence, indicating that the source supports the claim; and not enough information (NEI), indicating that the source lacks sufficient relevant information to assess the claim. Cell values denote numbers of claim–evidence pairs. SafeChecker increased correct classifications for the Inconclusive and Evidence classes while maintaining comparable performance for NEI, reducing three-class classification errors from 74 to 44. For binary unsupported-claim detection, Inconclusive and NEI were grouped as non-supporting evidence and treated as the positive class, whereas Evidence was treated as the negative class.

These Supplementary Notes provide the formal statements, assumptions, and proofs for the theoretical analysis summarized in the main text. Supplementary Note 1 fixes the definitions, notation, and standing conventions used throughout. Supplementary Notes 2–6 then address five theoretical questions in turn: the risk advantage of specialized agents over a monolithic one (Note 2), the bias and risk advantage of fusing heterogeneous sources over the best single source (Note 3), the distribution-free hallucination bound for the SafeChecker verification layer (Note 4), the termination and stability of the iterative refinement loop (Note 5), and the finite-sample generalization of a learned synthesis function to a deployment population (Note 6). Every theorem is a statement about an explicitly defined model of the corresponding MES design choice; a theoretical guarantee is not the same as empirical performance on benchmark data, nor the same as clinical validity, and the proofs below establish only the former under their stated hypotheses.

**Supplementary Note 1. Definitions, notation, and conventions**

The five analyses were developed in the mathematical setting most natural to each question and are complementary rather than nested: Notes 2 and 3 are statistical-decision-theoretic, Note 4 is distribution-free, Note 5 is a Lyapunov/stochastic-drift argument, and Note 6 is statistical-learning-theoretic. They share a common system-level interpretation, but each is stated in the framework native to its question rather than forced into a single formalism. This Note fixes the objects, symbols, and conventions that recur across the Notes and records once the assumptions that several results share, so that each subsequent Note can be read against a common vocabulary.

All random objects live on a common probability space $(\Omega, \mathcal{F}, \mathbb{P})$, with $\mathbb{E}$, $\mathrm{Var}$, and $\mathrm{Cov}$ the associated expectation, variance, and covariance. The true clinical estimand a query targets is $\theta^\star$, taken as a fixed unknown scalar $\theta^\star \in \mathbb{R}$ in Note 2 and as a fixed unknown vector $\theta^\star \in \mathbb{R}^d$ in Note 3. There are $K$ evidence sources, canonically $K = 3$, i.e., published literature, real-world data, and a biomedical knowledge graph; the source-specific estimator is written $\hat{\theta}_k$ (index $k$ in Note 2) or $\hat{\theta}_s$ (index $s$ in Note 3), each assumed square-integrable so that all biases, variances, covariances, and mean-squared-error risks below are finite. The bias of a source estimator is $b = \mathbb{E}[\hat{\theta}] - \theta^\star$ (scalar in Note 2, a vector in Note 3), and where it is useful to separate error that is common to all sources from error peculiar to one, we write $b = \beta + \delta$ with $\beta$ the shared component and $\delta$ the idiosyncratic component. Source estimators have (cross-)covariance $\Sigma$, symmetric and positive semidefinite.

Aggregation across sources is by convex weights $w$ in the probability simplex

$$\Delta^{K-1} = \left\{ w : w_k \geq 0, \sum_k w_k = 1 \right\},$$

encoding a weighted consensus on a common effect scale. The aggregated estimator is $T(w) = \sum_k w_k \, \hat{\theta}_k$, also written $\hat{\theta}_w$. The performance criterion in Notes 2 and 3 is mean-squared-error risk, $R(\cdot) = E \parallel \cdot - \theta^\star \parallel^2$. In Note 6, $R$ instead denotes the population or empirical risk of a learned synthesis function, $E\ell\{f(X), Y\}$; that notation is reintroduced there.

**Common-estimand applicability condition.** Notes 2 and 3 apply only to source outputs that estimate the same explicitly defined target quantity on a common scale. A mechanistic knowledge-graph relation, a trial effect and an observational association cannot be inserted into the same convex estimator merely because they concern the same clinical topic. Outputs that are not commensurable remain separate evidence components and may inform interpretation or weighting, but are not covered by the displayed MSE aggregation theorems.

The verification layer of Note 4 attaches to each candidate claim $c_i$ a latent truth label $T_i \in \{0,1\}$, with $T_i = 0$ denoting a false or unsupported claim, i.e., the *null event*. SafeChecker either accepts ($A_i = 1$) or does not, at a user-chosen tolerance $\alpha \in (0,1)$; the hallucination event is $H_i = \{A_i = 1,\ T_i = 0\}$, a false claim accepted into the report. These four symbols $(T_i, \alpha, A_i, H_i)$ are used with this meaning wherever verification is discussed, including the composition statement in Note 5.

To avoid notational ambiguity across Notes, several symbols are localized. The symbol $R$ denotes risk throughout. In Note 4, the SafeChecker nonconformity and contradiction scores are $s_i$ and $\kappa_i$, whereas $K$ denotes the number of sources. The letter $\rho$ is section-localized: pairwise correlation in Note 2, distance $\rho^\star$ from the origin to the bias hull in Note 3, a conformal rank in Note 4 and the geometric drift or contraction factor in Note 5. The letter $b$ denotes source bias in Notes 2–3 and an additive drift allowance in Note 5; it is not necessarily a lower bound or limiting residual. Metric weights in Note 5 are $\mu_\bullet$, so $\lambda_w$ can denote the joint optimal risk in Notes 3 and 6. The letter $\delta$ denotes an idiosyncratic bias when subscripted in Notes 2–3 and a failure probability in Note 6. These conventions are not substantive assumptions.

The hypotheses common to several Notes are deliberately limited: all objects live on a common probability space, estimators lie in $L^2$ wherever MSE risk is used, source weights lie in the simplex and source outputs are aggregated only when they target a common estimand on a common scale. Each result then rests on its own substantive hypothesis: a joint capacity-interference model for the monolithic estimator in Note 2; a directional-diversity condition on source biases in Note 3; null exchangeability in Note 4; finite-progress conditions and, separately, a stochastic-drift condition in Note 5; and bounded loss, measurability and independent source samples in Note 6. These are model assumptions, not empirical consequences of the architecture.

**Supplementary Note 2. Multi-agent versus single-agent estimation risk**

This Note asks whether decomposing evidence synthesis into specialized retrieval agents followed by optimal aggregation yields an estimator of a clinical effect whose risk is no larger, i.e., and under diversity, strictly smaller, than that of a single monolithic agent performing the same task end-to-end. We fix a clinical query with scalar estimand $\theta^\star \in \mathbb{R}$ (a vector extension is given at the end of the Note). There are $K \geq 2$ evidence sources; source $k$ admits a specialist estimator $\hat{\theta}_k \in L^2(\mathbb{P})$ with bias $b_k := \mathbb{E}[\hat{\theta}_k] - \theta^\star$, variance $\sigma_k^2 := \mathrm{Var}(\hat{\theta}_k)$, covariance matrix $\Sigma = (\Sigma_{k\ell})$ with $\Sigma_{kk} = \sigma_k^2$, and $b := (b_1, \dots, b_K)^\top$. An aggregator forms $T(w) := \sum_k w_k \hat{\theta}_k$ for $w \in \Delta^{K-1}$, and the performance criterion is the mean-squared-error risk $R(T) := \mathbb{E}[(T - \theta^\star)^2]$.

Every bound below is read through the classical bias–variance[1] decomposition of mean-squared error: for any $T \in L^2(\mathbb{P})$ and fixed $\theta^\star$, writing $\mu = \mathbb{E}[T]$ and expanding $R(T) = \mathbb{E}\left[\left((T-\mu) + (\mu - \theta^\star)\right)^2\right]$, the cross term vanishes and

$$R(T) = \mathrm{Bias}(T)^2 + \mathrm{Var}(T), \qquad \mathrm{Bias}(T) = \mathbb{E}[T] - \theta^\star.$$

Applied to the aggregated estimator, linearity gives $\mathrm{Bias}\big(T(w)\big) = w^\top b$ and $\mathrm{Var}\big(T(w)\big) = w^\top \Sigma w \geq 0$, so that

$$R\big(T(w)\big) = (w^\top b)^2 + w^\top \Sigma\, w. \tag{2.1}$$

Both terms are convex functionals of the weights, and the consensus improves on the typical source precisely through this convexity. By Jensen's inequality for the map $x \mapsto x^2$ against the probability weights $w$, $(w^\top b)^2 \leq \sum_k w_k\, b_k^2$, with equality only when all biases carrying positive weight are equal; and by Cauchy–Schwarz in $L^2$, $|\Sigma_{k\ell}| \leq \sigma_k \sigma_\ell$, so $w^\top \Sigma w \leq (\sum_k w_k\, \sigma_k)^2 \leq \sum_k w_k\, \sigma_k^2$, with the first equality only when all weighted sources are perfectly correlated and the second only when their standard deviations agree. Substituting these into the above equation gives the assumption-free bound

$$R\big(T(w)\big) \leq \sum_k w_k\, (b_k^2 + \sigma_k^2) = \sum_k w_k\, R\big(\hat{\theta}_k\big),$$

Thus, the risk of any weighted consensus never exceeds the weighted average of the source risks. A strict variance gap is a standard within-vector diversification gain. The architecture-level comparison below is a distinct result and additionally requires the stated joint PSD covariance-inflation model.

We model the vector of monolithic source sub-estimators as

$$\hat{\boldsymbol{\theta}}_0 = \hat{\boldsymbol{\theta}} + \boldsymbol{\xi}, \qquad \hat{\boldsymbol{\theta}} = (\hat{\theta}_1, \dots, \hat{\theta}_K)^\top.$$

It is important to distinguish a marginal condition from the stronger joint condition needed for a covariance-matrix comparison.

**Lemma 2.1 (Marginal and joint effects of capacity-interference noise).** Let all components be square-integrable.

(a) If $E[\xi_k \mid \hat{\theta}_k] = 0$ for a given $k$, then

$$E[\hat{\theta}_{0,k}] = E[\hat{\theta}_k], \qquad \mathrm{Var}(\hat{\theta}_{0,k}) = \mathrm{Var}(\hat{\theta}_k) + \mathrm{Var}(\xi_k).$$

Thus, the marginal bias is unchanged and the marginal variance is weakly increased.

(b) If the stronger joint condition

$$E[\boldsymbol{\xi} \mid \widehat{\boldsymbol{\theta}}] = \mathbf{0} \tag{J}$$

holds, then, writing $\Sigma = \mathrm{Cov}(\widehat{\boldsymbol{\theta}})$, $\Xi = \mathrm{Cov}(\boldsymbol{\xi})$ and $\Sigma_0 = \mathrm{Cov}(\widehat{\boldsymbol{\theta}}_0)$,

$$\mathbf{b}_0 = \mathbf{b}, \qquad \mathrm{Cov}(\widehat{\boldsymbol{\theta}}, \boldsymbol{\xi}) = 0, \qquad \Sigma_0 = \Sigma + \Xi \succcurlyeq \Sigma.$$

**Proof.** For part (a),

$$E[\xi_k] = E\{E[\xi_k \mid \hat{\theta}_k]\} = 0$$

and

$$\mathrm{Cov}(\hat{\theta}_k, \xi_k) = E\big[(\hat{\theta}_k - E\hat{\theta}_k)E(\xi_k \mid \hat{\theta}_k)\big] = 0.$$

Expanding the variance of $\hat{\theta}_{0,k} = \hat{\theta}_k + \xi_k$ proves part (a). For part (b), iterated expectation gives $E[\boldsymbol{\xi}] = 0$ and

$$\mathrm{Cov}(\widehat{\boldsymbol{\theta}}, \boldsymbol{\xi}) = E\big[(\widehat{\boldsymbol{\theta}} - E\widehat{\boldsymbol{\theta}})E(\boldsymbol{\xi}^\top \mid \widehat{\boldsymbol{\theta}})\big] = 0.$$

Its transpose is also zero. Therefore

$$\Sigma_0 = \mathrm{Cov}(\widehat{\boldsymbol{\theta}} + \boldsymbol{\xi}) = \Sigma + \Xi.$$

Because $\Xi$ is a covariance matrix, $\Xi \succcurlyeq 0$. ■

Part (a) is a marginal statement and does not imply part (b). Theorem 2.1 uses (J), or equivalently takes $\mathbf{b}_0 = \mathbf{b}$ and $\Sigma_0 = \Sigma + \Xi$, $\Xi \succcurlyeq 0$, as its explicit model assumption. This is a sufficient theoretical model, not an empirical consequence of specialization.

$$R^{\mathrm{SA}} := \min_{v \in \Delta^{K-1}} [(v^\top b_0)^2 + v^\top \Sigma_0 v], \qquad R^{\mathrm{MA}} := \min_{w \in \Delta^{K-1}} [(w^\top b)^2 + w^\top \Sigma w], \tag{2.2}$$

both minima being attained since the objective is continuous and the simplex compact. The main result now follows by a one-line feasibility comparison.

**Theorem 2.1 (Conditional risk dominance under joint covariance inflation).** Suppose that the specialist and monolithic sub-estimator vectors have the same bias vector, $\mathbf{b}_0 = \mathbf{b}$, and covariance matrices satisfying

$$\Sigma_0 = \Sigma + \Xi, \qquad \Xi \succcurlyeq 0. \tag{C}$$

Define

$$R_{\mathrm{SA}} = \min_{v \in \Delta^{K-1}} \{(v^\top \mathbf{b})^2 + v^\top \Sigma_0 v\},$$

and

$$R_{\mathrm{MA}} = \min_{w \in \Delta^{K-1}} \{(w^\top \mathbf{b})^2 + w^\top \Sigma w\}.$$

Then $R_{\mathrm{MA}} \le R_{\mathrm{SA}}$. If $v^\star$ is any minimizer of the single-agent problem and $v^{\star\top}\Xi v^\star > 0$, then $R_{\mathrm{MA}} < R_{\mathrm{SA}}$. Condition (C) follows from Lemma 2.1(b) under the sufficient joint condition $E[\boldsymbol{\xi} \mid \widehat{\boldsymbol{\theta}}] = \mathbf{0}$.

**Proof.** The objectives are continuous and the simplex is compact, so minimizers exist. Let $v^\star$ minimize the single-agent objective. It is feasible for the multi-agent problem, and hence

$$R_{\mathrm{MA}} \le (v^{\star\top}\mathbf{b})^2 + v^{\star\top}\Sigma v^\star.$$

Moreover,

$$(v^{\star\top}\mathbf{b})^2 + v^{\star\top}\Sigma v^\star = (v^{\star\top}\mathbf{b})^2 + v^{\star\top}\Sigma_0 v^\star - v^{\star\top}\Xi v^\star = R_{\mathrm{SA}} - v^{\star\top}\Xi v^\star \le R_{\mathrm{SA}}.$$

Strictness follows when the displayed quadratic form is positive. ■

At the single-agent optimizer $v^\star$, removing the modeled interference lowers the displayed comparator risk by exactly $v^{\star\top}\Xi v^\star$. Because the multi-agent problem reoptimizes over all feasible weights, the optimized gap satisfies

$$R_{\mathrm{SA}} - R_{\mathrm{MA}} \ge v^{\star\top}\Xi v^\star;$$

it need not equal this quadratic form. The theorem makes no claim of bias reduction because the bias vectors are held equal. Bias geometry is treated separately below, and the uncorrelated special case gives an explicit diversification calculation.

**Corollary 2.2 (Inverse-variance diversification).** If the source estimators are mutually uncorrelated, $\Sigma = \mathrm{diag}(\sigma_1^2, \ldots, \sigma_K^2)$ with $\sigma_k^2 > 0$, and share no common bias direction (take $b = 0$), then the optimal weight is the inverse-variance weight and

$$w_k^{\mathrm{IV}} = \frac{1/\sigma_k^2}{\sum_\ell 1/\sigma_\ell^2}, \qquad R^{\mathrm{MA}} = (\sum_k 1/\sigma_k^2)^{-1} \le \min_k \sigma_k^2,$$

strict once $K \ge 2$ with all $\sigma_k^2 < \infty$.

*Proof.* With $b = 0$ and diagonal $\Sigma$ the objective is $\sum_k w_k^2\, \sigma_k^2$; Lagrange stationarity $2w_k\sigma_k^2 = \lambda$ gives $w_k \propto 1/\sigma_k^2$. The objective is strictly convex on the affine constraint, so this is the unique global minimum, and substituting yields $(\sum_k \sigma_k^{-2})^{-1} \le \min_k \sigma_k^2$, strict for $K \ge 2$.

For correlated sources, the gap between the weighted-average variance and the achieved variance is

$$\sum_k w_k\, \sigma_k^2 - w^\top \Sigma w = \frac{1}{2}\sum_{k,\ell} w_k\, w_\ell (\sigma_k - \sigma_\ell)^2 + 2\sum_{k<\ell} w_k\, w_\ell \sigma_k \sigma_\ell (1 - \rho_{k\ell}) \ge 0.$$

The gap is strictly positive exactly when at least one non-negative summand involving sources with positive weights is positive. A vertex weight has zero diversification gap; for a multi-source positive-weight support, zero gap requires equiprecision and perfect pairwise correlation on that support.

The bias comparison is governed by a shifted convex-hull geometry. Write $b_k = \beta + \delta_k$, where $\beta$ is a common offset and $\delta_k$ is source-specific. Then

$$\mathrm{Bias}\{T(w)\} = \beta + w^\top \delta, \qquad w \in \Delta^{K-1}.$$

The common offset need not be a lower bound on total absolute bias because an admissible idiosyncratic mixture may partially or exactly cancel it.

**Corollary 2.3 (Geometry of a shared offset and idiosyncratic biases).** Suppose $b_k = \beta + \delta_k$ and let $\mathcal{D} = \mathrm{conv}\{\delta_1, \ldots, \delta_K\}$.
(a) For every $w \in \Delta^{K-1}$,

$$||\beta| - |w^\top \delta|| \le |\beta + w^\top \delta| \le |\beta| + |w^\top \delta|,$$

and

$$\min_{w \in \Delta^{K-1}} |\beta + w^\top \delta| = \operatorname{dist}(-\beta, \mathcal{D}). \tag{2.3}$$

The total bias can be zero if and only if $-\beta \in \mathcal{D}$; $|\beta|$ is not a universal lower bound.

(b) For every $w \in \Delta^{K-1}$,

$$(w^\top \delta)^2 \le \sum_{k=1}^{K} w_k\, \delta_k^2.$$

If $\min_k \delta_k < 0 < \max_k \delta_k$ , some convex mixture has $w^\top \delta = 0$ . This cancels the idiosyncratic component but minimizes total bias only when zero is a closest point in $\mathcal{D}$ to $-\beta$, for example when $\beta = 0$.

(c) Let a comparator have total bias $\beta + \delta_0$ with $\delta_0 \in \mathcal{D}$, and let

$$w^\dagger \in \arg\min_{w \in \Delta^{K-1}} |\beta + w^\top \delta|.$$

Then

$$|\beta + w^{\dagger\top} \delta| \le |\beta + \delta_0|,$$

strictly if and only if $\delta_0$ is not a closest point in $\mathcal{D}$ to $-\beta$.

**Proof.** The inequalities in part (a) are the triangle and reverse-triangle inequalities. As $w$ ranges over the simplex, $w^\top \delta$ ranges exactly over $\mathcal{D}$, giving the distance formula and the zero-bias equivalence. Part (b) is Jensen's inequality. In the scalar case, $\mathcal{D} = [\min_k \delta_k, \max_k \delta_k]$, so strict sign diversity places zero in the interval. For part (c), $\delta_0 \in \mathcal{D}$ is feasible in (2.3); optimality of $w^\dagger$ gives the comparison, with equality exactly at another closest point. ■

Corollary 2.3 is an oracle geometry, not a guarantee that MES reduces total bias. Distinct source mechanisms do not determine the signs or magnitudes of $\delta_k$, and SafeChecker does not observe these bias components. Total-bias improvement occurs only when the shifted bias hull is closer to zero than the comparator, as stated in part (c).

**Proposition 2.4 (No-gain boundary).** If simultaneously (i) $\Xi = 0$ (no dilution), (ii) all $\rho_{k\ell} = 1$ and all $\sigma_k^2 = \sigma^2$ (redundant sources), and (iii) $\delta_k = 0$ for all $k$ (common bias only), then for every $w$, $T(w)$ has bias $\beta$ and variance $\sigma^2$, so $R^{\mathrm{MA}} = R^{\mathrm{SA}} = \beta^2 + \sigma^2$ and there is no multi-agent advantage.

*Proof.* Direct substitution into the displayed risk identity: the bias is $\beta \sum_k w_k = \beta$ and the variance $\sigma^2 (\mathbf{1}^\top w)^2 = \sigma^2$, both weight-independent.

Proposition 2.4 identifies one no-gain boundary but is not an if-and-only-if characterization. Theorem 2.1 supplies a sufficient architecture-level comparison under joint PSD covariance inflation, whereas Corollary 2.3 describes a separate oracle bias geometry. Neither result establishes that these conditions hold empirically in MES.

Random-effects modeling can be incorporated only after specifying the augmented joint covariance matrix; the preceding inequalities then apply if that matrix satisfies the same positive-semidefinite and cross-covariance conditions. Optional reliability-based reweighting does not enlarge the simplex: deleting or constraining weights can increase the minimum risk. Such reweighting must be evaluated through its full bias–variance or generalization certificate.

For a vector estimand $\theta^\star \in \mathbb{R}^d$, stack source estimators as

$$\widehat{\Theta} = (\hat{\theta}_1^\top, \dots, \hat{\theta}_K^\top)^\top \in \mathbb{R}^{Kd},$$

define $A_w = w^\top \otimes I_d$, and let $b \in \mathbb{R}^{Kd}$ and $\Sigma \in \mathbb{R}^{Kd \times Kd}$ be the stacked bias and covariance. Then

$$R\{A_w\widehat{\Theta}\} = \| A_w b \|_2^2 + \mathrm{tr}(A_w \Sigma A_w^\top).$$

If $\Sigma_0 = \Sigma + \Xi$, $\Xi \succcurlyeq 0$, and the stacked biases agree, Theorem 2.1 carries over with non-negative gap

$$\mathrm{tr}(A_v \Xi A_v^\top) \geq 0.$$

**Supplementary Note 3. Multi-source versus single-source bias and risk**

This Note asks whether fusing heterogeneous evidence sources by convex combination produces an estimator no more biased and no higher in risk than the best single source, and strictly better under a checkable directional-diversity condition. We work in $\mathbb{R}^d$, with $d = 1$ the canonical single-effect case. The estimand is $\theta^\star \in \mathbb{R}^d$; sources $s \in \{1, \ldots, K\}$ have estimators $\hat{\theta}_s \in L^2(\mathbb{P}; \mathbb{R}^d)$ with bias vectors $b_s := \mathbb{E}[\hat{\theta}_s] - \theta^\star$ and covariance blocks $\Sigma_{st} := \mathrm{Cov}(\hat{\theta}_s, \hat{\theta}_t)$. The fused estimator is $\hat{\theta}_w := \sum_s w_s \hat{\theta}_s$ for $w \in \Delta^{K-1}$, with fused bias $B(w) := \sum_s w_s b_s$ and risk $R(T) := \mathbb{E} \parallel T - \theta^\star \parallel^2$.

As in Note 2 the argument begins from the bias–variance decomposition, now in vector form. For $T \in L^2(\mathbb{P}; \mathbb{R}^d)$, expanding about $\mu = \mathbb{E}[T]$ and discarding the vanishing cross term gives $R(T) =\parallel \mathrm{Bias}(T) \parallel^2 + \mathrm{tr}\mathrm{Cov}(T)$, so that

$$R(\hat{\theta}_w) =\parallel \sum_s w_s b_s \parallel^2 + \sum_{s,t} w_s w_t \mathrm{tr}\Sigma_{st}.$$

The fused bias $B(w) = \sum_s w_s b_s$ ranges, as $w$ sweeps the simplex, over exactly the convex hull $\mathrm{conv}\{b_1, \ldots, b_K\}$ of the source bias vectors, so the smallest bias fusion can achieve is governed entirely by the position of the origin relative to that hull. Writing $\rho^\star$ for the distance from the origin to the bias hull (a Note-3-local meaning of $\rho$), define the minimal achievable bias norm

$$\rho^\star := \min_{w \in \Delta^{K-1}} \parallel B(w) \parallel = \mathrm{dist}(0, \mathrm{conv}\{b_1, \ldots, b_K\}),$$

attained at some $w^\star$ by compactness, and let $m := \min_s \parallel b_s \parallel$ be the best single-source bias, attained at $i^\star$. The central bias result compares these two.

**Theorem 3.1 (Multi-source bias dominance).** (a) Always $\rho^\star \leq m$. (b) If some source $j$ satisfies the directional-diversity condition

$$\langle b_{i^\star}, b_j \rangle < \parallel b_{i^\star} \parallel^2 \qquad \text{(D)}$$

(equivalently, $\langle b_{i^\star}, b_j - b_{i^\star} \rangle < 0$), then $\rho^\star < m$; in particular, (D) holds whenever $\langle b_{i^\star}, b_j \rangle \leq 0$. (c) In the scalar case $d = 1$,

$$\rho^\star = \mathrm{dist}\left(0, [\min_s b_s, \max_s b_s]\right).$$

Hence $\rho^\star = 0$ if and only if $\min_s b_s \leq 0 \leq \max_s b_s$. Strict improvement $\rho^\star < m$ occurs if and only if $m > 0$ and the interval contains zero.

*Proof.* For (a), $w = e_{i^\star}$ is feasible and gives $\parallel B(e_{i^\star}) \parallel = m$, so $\rho^\star \leq m$. For (b), along $w(t) = (1-t)e_{i^\star} + te_j$ we have $g(t) := \parallel B(w(t)) \parallel^2 = \parallel b_{i^\star} \parallel^2 + 2t\langle b_{i^\star}, b_j - b_{i^\star} \rangle + t^2 \parallel b_j - b_{i^\star} \parallel^2$, so $g'(0) = 2(\langle b_{i^\star}, b_j \rangle - \parallel b_{i^\star} \parallel^2) < 0$ by (D); hence some $t_0 \in (0,1]$ gives $\parallel B(w(t_0)) \parallel < m$, so $\rho^\star < m$. For (c), in $d = 1$ the hull $\mathrm{conv}\{b_s\} = [\min_s b_s, \max_s b_s]$ has distance to $0$ equal to $0$ iff $0$ lies in the interval, else the nearest endpoint has norm $\min_s |b_s|$.

Condition (D) is a sufficient population-level directional condition. Structurally different evidence mechanisms may make diverse error directions plausible but do not imply (D); the true bias vectors are unobserved and the theorem does not certify that MES satisfies it.

**Corollary 3.2 (Exact cancellation).**

$$\rho^\star = 0 \quad \Leftrightarrow \quad 0 \in \mathrm{conv}\{b_1, \ldots, b_K\}.$$

If zero lies in the relative interior of this hull, it admits a representation with strictly positive weights on all listed sources. The fused estimator is then unbiased and is strictly less biased than every source provided $\min_s \| b_s \|_2 > 0$. Exact cancellation may occur with fewer than $d+1$ sources in a lower-dimensional affine subspace; two opposing vectors suffice in any ambient dimension. Containing zero in the full-dimensional interior of a hull in $\mathbb{R}^d$, however, requires at least $d+1$ affinely spanning sources.

**Proof.** The equivalence follows from the definition of $\rho^\star$ as the distance to the compact bias hull. A relative-interior point of a finite convex hull has a representation assigning positive mass to all listed points. Strict comparison requires that no source is itself unbiased. The dimensional claim follows from affine dimension; Carathéodory's theorem does not impose $d+1$ points on lower-dimensional cancellation. ■

Just as in Note 2, fusion cannot touch error that every source shares; the next proposition records this floor in vector form.

**Proposition 3.3 (Shared bias is irreducible).** Writing $b_s = \beta + \delta_s$ with $\beta \in \mathbb{R}^d$ the shared component, for every $w$, $\| B(w) \| \geq \| \beta \| - \| \sum_s w_s \delta_s \|$. If all $\delta_s$ are orthogonal to $\beta$, then $\langle B(w), \beta \rangle = \| \beta \|^2$ for all $w$, so $\| B(w) \| \geq \| \beta \|$: no convex fusion reduces bias below the shared component.

**Proof.** The first bound is the reverse triangle inequality. In the orthogonal case,

$$\langle B(w), \beta \rangle = \| \beta \|^2 + \sum_s w_s \langle \delta_s, \beta \rangle = \| \beta \|^2.$$

If $\beta = 0$, the claimed lower bound is immediate. If $\| \beta \| > 0$, Cauchy–Schwarz gives $\| \beta \|^2 \leq \| B(w) \| \, \| \beta \|$, and division by $\| \beta \|$ completes the proof.

Bias is only half the story, since the risk depends on variance as well, and the risk comparison has the same shape as the bias comparison. Define $R^{\mathrm{MA}} := \min_w R(\hat{\theta}_w)$ and the best single-source risk $R^{\mathrm{SS}} := \min_s R(\hat{\theta}_s) = \min_s [\| b_s \|^2 + \mathrm{tr}\Sigma_s]$.

**Theorem 3.4 (Multi-source risk dominance).** (a) Always $R^{\mathrm{MA}} \leq R^{\mathrm{SS}}$. (b) Strict dominance $R^{\mathrm{MA}} < R^{\mathrm{SS}}$ holds if, with $s^\star \in \mathrm{argmin}_s R(\hat{\theta}_s)$, some source $j$ satisfies

$$\langle b_{s^\star}, b_j - b_{s^\star} \rangle + \left(\mathrm{tr}\Sigma_{s^\star j} - \mathrm{tr}\Sigma_{s^\star}\right) < 0. \tag{R}$$

(c) If sources are uncorrelated and unbiased ($\Sigma_{st} = 0$ for $s \neq t$, $b_s = 0$), then with $v_s := \mathrm{tr}\Sigma_s > 0$ the risk-optimal weights are inverse-variance, $w_s^\dagger = (1/v_s) / \sum_\ell (1/v_\ell)$, and $R^{\mathrm{MA}} = (\sum_s 1/v_s)^{-1} \leq \min_s v_s = R^{\mathrm{SS}}$, strict for $K \geq 2$.

*Proof.* For (a), $w = e_{s^\star}$ is feasible with value $R^{\mathrm{SS}}$. For (b), along $w(t) = (1-t)e_{s^\star} + t e_j$, $\frac{d}{dt}|_0 R(\hat{\theta}_{w(t)}) = 2\langle b_{s^\star}, b_j - b_{s^\star} \rangle + 2\left(\mathrm{tr}\Sigma_{s^\star j} - \mathrm{tr}\Sigma_{s^\star}\right) < 0$ under (R), a feasible decrease. For (c), minimize $\sum_s w_s^2 v_s$ over the simplex (strictly convex), Lagrange gives $w_s \propto 1/v_s$, and substitution yields the stated value.

The weights that minimize bias and those that minimize risk are generally distinct, and the two claims concern different fused estimators. The bias-minimizing weight $w^\star$ of Theorem 3.1 and the risk-minimizing weight $w^\dagger$ of Theorem 3.4 can differ because reducing bias may place substantial weight on noisy sources. A narrower local implication follows from (R): from a best-risk vertex $s^\star$, a direction $j$ that decreases squared bias also decreases risk whenever

$$\mathrm{tr}\Sigma_{s^\star j} \leq \mathrm{tr}\Sigma_{s^\star}.$$

Independence is one sufficient case because the cross-covariance term is zero. This local implication does not make the global bias and risk objectives, or their optimizers, coincide.

The general optimizers in Theorems 3.1 and 3.4 are oracle quantities. The bias-optimal weight depends on the unknown $b_s$, and the general risk-optimal weight depends on both the unknown biases and the full covariance blocks $\Sigma_{st}$. Covariance estimation alone therefore does not realize the general oracle risk minimum. The directly computable result is Theorem 3.4(c), where unbiased, mutually uncorrelated sources yield inverse-variance weights. In deployment, MES uses uncertainty, provenance, study quality and cross-source consistency as heuristic proxies; the theory motivates these inputs but does not prove that the resulting weights attain either oracle frontier.

**Supplementary Note 4. Distribution-free control of hallucination risk by SafeChecker**

This Note asks what the SafeChecker verification layer can guarantee about the rate at which false or unsupported claims are accepted into a report, at a finite, user-chosen level $\alpha$, without any distributional model of the underlying score. Because the verification layer's notation overlaps with the estimation notation used in Notes 2 and 3, several symbols are localized to this Note. The nonconformity score of claim $c_i$ is $\mathsf{s}_i$, with larger values indicating lower trustworthiness; the contradiction score is κi, summarizing negation and contrapositive probing. The provenance indicator is $G_i \in [0,1]$, the aggregate support is $S_i = \sum_j \alpha_{ij} q_j$, the internal-consistency summary is $V_i \in [0,1]$, the nonconformity score of calibration item $Z_k$ is $\mathsf{s}_k^{\text{cal}}$, the rank of $\mathsf{s}_i$ in the augmented sample is $\rho_i \in \{1, \dots, N+1\}$, the acceptance threshold is a low order statistic $\tau_\alpha$, and $\ell_\alpha = \lfloor (N+1)\alpha \rfloor$ is the calibration index. The quantities $\alpha, \beta, q$ denote the false-acceptance level, false-rejection level, and FDR level, respectively; $F_0$ denotes the false-claim score CDF; and $\eta_\beta, \kappa$ appear in the detectability route. Outside this Note, these symbols revert to their global meanings; the truth label $T_i \in \{0,1\}$, level $\alpha$, acceptance $A_i$, and hallucination event $H_i = \{A_i = 1, T_i = 0\}$ are the shared symbols of Note 1.

For a report with candidate claims $\{c_1, \dots, c_m\}$ over an evidence pool $\{e_1, \dots, e_n\}$, SafeChecker computes per claim the provenance indicator $G_i$, the support $S_i = \sum_j \alpha_{ij} q_j$ (with $\alpha_{ij} \in [0,1]$ relevance and $q_j \in [0,1]$ quality), the contradiction $\kappa_i \in [0, \infty)$, and the uncertainty $V_i \in [0,1]$, and aggregates them into a single scalar nonconformity score $\mathsf{s}_i = r(G_i, S_i, \kappa_i, V_i) \in \mathbb{R}$ with $r$ measurable and oriented so that larger means less trustworthy. A canonical monotone instance is $\mathsf{s}_i = \gamma_1(1 - \tilde{S}_i) + \gamma_2 \tilde{\kappa}_i + \gamma_3(1 - G_i) + \gamma_4(1 - V_i)$ with $\gamma_\ell \geq 0$ and min–max normalized inputs, but no parametric form is required; the results use only measurability and orientation. The latent label is $T_i \in \{0,1\}$, where $T_i = 1$ if $c_i$ is true or adequately supported and $T_i = 0$ if false, unsupported, or materially misleading, and accepting a $T_i = 0$ claim is a hallucination passing the verifier.

The mechanism is calibrated from a set of known-false claims, following the split-conformal construction.[2] Let $\mathcal{D}_{\text{cal}}^0 = \{Z_1, \dots, Z_N\}$ with $T_k^{\text{cal}} = 0$ for all $k$, each scored $\mathsf{s}_k^{\text{cal}} := r(Z_k)$ by the same pipeline as a test claim. Because a low score means apparently trustworthy, a false claim is dangerous when its score is small, so we calibrate from the lower tail: with order statistics $\mathsf{s}_{(1)}^{\text{cal}} \leq \cdots \leq \mathsf{s}_{(N)}^{\text{cal}}$ and level $\alpha \in (0,1)$, set the acceptance threshold $\tau_\alpha := \mathsf{s}_{(\ell_\alpha)}^{\text{cal}}$ with $\ell_\alpha := \lfloor (N+1)\alpha \rfloor$, and accept iff the score is strictly below it, $A_i := \mathbf{1}\{\mathsf{s}_i < \tau_\alpha\}$. When $\ell_\alpha = 0$, i.e. $\alpha < 1/(N+1)$, we set $\tau_\alpha := -\infty$ so $A_i \equiv 0$: the calibration sample is too small to certify anything at that level and SafeChecker abstains, an honest conservative default. Such a false-claim set is realizable in MES, since SafeChecker's negation and contrapositive probing already manufactures known-false claims, logical negations, wrong-sign effects, fabricated or retracted citations, supplemented by red-team and expert-adjudicated false claims.

The guarantee rests on a single probabilistic assumption together with three structural ones. The substantive hypothesis is null-exchangeability (A1): for a test claim on $\{T_i = 0\}$, the augmented sequence $(\mathsf{s}_1^{\text{cal}}, \dots, \mathsf{s}_N^{\text{cal}}, \mathsf{s}_i)$ is exchangeable, its joint law invariant under all $(N+1)!$ coordinate permutations. This is coherent because calibration and test scores are now drawn from the same false-claim population; it holds in particular if they are i.i.d. from a common null law $F_0$, but A1 is strictly weaker and permits symmetric dependence, and it is the only load-bearing probabilistic assumption. The structural conditions are monotone orientation (A2: $r$ oriented so larger $\mathsf{s}$ means lower trustworthiness, with the one-sided rule $A_i = \mathbf{1}\{\mathsf{s}_i < \tau_\alpha\}$), measurability (A3: $r$ measurable, all scores, ranks, and events well-defined), and a generic no-ties condition (A4: $\mathbb{P}\big(\mathrm{s}_i = \mathrm{s}_k^{\mathrm{cal}} \mid T_i = 0\big) = 0$), the last presentational only and discharged below by randomized tie-breaking. The analysis assumes no parametric form for $\mathrm{s}$, no independence beyond the symmetric dependence A1 permits, no knowledge of $F_0$, no calibration of component false-positive rates, and nothing about the true-class score law; the guarantee is distribution-free in $F_0$.

The analysis follows from a single rank-symmetry fact.
**Lemma 4.1 (Uniform rank of the test score under the null).** Under A1, A3, A4, for a test claim with $T_i = 0$ define $\rho_i := 1 + \sum_{k=1}^{N} \mathbf{1}\{\mathrm{s}_k^{\mathrm{cal}} < \mathrm{s}_i\} \in \{1, \dots, N+1\}$. Then $\rho_i$ is uniform on $\{1, \dots, N+1\}$, $\mathbb{P}(\rho_i = r \mid T_i = 0) = 1/N+1$, and for every integer $k \in \{0, \dots, N+1\}$, $\mathbb{P}(\rho_i \le k \mid T_i = 0) = k/N+1$.

*Proof.* Condition on $T_i = 0$. By A4 the $N+1$ scores are a.s. distinct, so the rank $\rho_i$ is well-defined and takes each value in $\{1, \dots, N+1\}$ on disjoint events partitioning the space. By A1 the joint law is invariant under the symmetric group $\mathfrak{S}_{N+1}$; the rank of the last coordinate is a function of the configuration, and for any ranks $r, r'$ a transposition carries "last coordinate has rank $r$" to "rank $r'$", so exchangeability makes these equiprobable and each of the $N+1$ partitioning events has probability $1/(N+1)$. The cumulative statement sums $k$ disjoint atoms, trivial at $k = 0$.
Here A4 supplies a well-defined strict rank, A1 supplies equiprobability, and A3 ensures measurability; this is the single engine behind every bound below. The per-claim guarantee follows by reading the acceptance event through the rank.

**Theorem 4.1 (Exact per-claim false-acceptance bound).** Assume A1–A4, with $\tau_\alpha = \mathrm{s}_{(\ell_\alpha)}^{\mathrm{cal}}$ and $A_i = \mathbf{1}\{\mathrm{s}_i < \tau_\alpha\}$. Then for every test claim with $T_i = 0$,

$$\mathbb{P}(A_i = 1 \mid T_i = 0) \;\le\; \frac{\lfloor (N+1)\alpha \rfloor}{N+1} \;\le\; \alpha,$$

and consequently $\mathbb{P}(H_i) = \mathbb{P}(A_i = 1, T_i = 0) \le \alpha\, \mathbb{P}(T_i = 0) \le \alpha$.

*Proof.* Condition on $T_i = 0$. If $\mathrm{s}_i < \mathrm{s}_{(\ell_\alpha)}^{\mathrm{cal}}$ then $\mathrm{s}_i$ is strictly below at least $N - \ell_\alpha + 1$ calibration scores, hence strictly above at most $\ell_\alpha - 1$, so $\rho_i \le \ell_\alpha$. Thus $\{\mathrm{s}_i < \tau_\alpha\} \subseteq \{\rho_i \le \ell_\alpha\}$, and by Lemma 4.1, $\mathbb{P}(A_i = 1 \mid T_i = 0) \le \ell_\alpha/(N+1) = \lfloor (N+1)\alpha \rfloor/(N+1) \le \alpha$, the last step since $\lfloor x \rfloor \le x$. The unconditional bound follows from $\mathbb{P}(H_i) = \mathbb{P}(A_i = 1 \mid T_i = 0)\, \mathbb{P}(T_i = 0)$ and $\mathbb{P}(T_i = 0) \le 1$.

Every inequality tightens in the safe direction, the set inclusion can only lower the acceptance probability, and $\lfloor (N+1)\alpha \rfloor/(N+1) \le \alpha$, so the bound is a genuine ceiling on hallucination acceptance, converting an uncalibrated score into a rule with a guaranteed rate using only rank symmetry, where an accept-all verifier would have rate $1$ and an arbitrary fixed threshold an uncontrolled rate $F_0$(threshold). The bound is also essentially exact: if the augmented scores are i.i.d. from a continuous $F_0$ (a special case of A1), then $\mathbb{P}(A_i = 1 \mid T_i = 0) = \ell_\alpha/(N+1)$ with equality and $\lfloor (N+1)\alpha \rfloor/(N+1) \uparrow \alpha$ as $N \to \infty$, the gap to $\alpha$ being the unavoidable finite-sample rounding $O(1/N)$ (the inclusion becomes an a.s. equality and Lemma 4.1 gives the rank probability exactly, with $(N+1)\alpha - 1 < \lfloor (N+1)\alpha \rfloor \le (N+1)\alpha$). The no-ties assumption A4 is moreover only presentational: with positive-probability ties, attach i.i.d. $U_0, \dots, U_N \sim \mathrm{Unif}(0,1)$ to the scores and break ties by $U$-value, after which the augmented pairs remain exchangeable under A1 and are a.s. distinct, so Lemma 4.1 and Theorem 4.1 hold verbatim for the randomized rank. The same rule is conveniently packaged as a conformal p-value.[3]

**Proposition 4.2 (Conformal p-value / super-uniformity).** Define $\tilde{p}_i := \rho_i/(N+1) = \big(1 +$

$\sum_k \mathbf{1}\{s_k^{\mathrm{cal}} < s_i\})/(N+1)$. Then $\tilde{p}_i$ is super-uniform on the null, $\mathbb{P}(\tilde{p}_i \le \alpha \mid T_i = 0) \le \alpha$ for all $\alpha \in (0,1)$, and accepting iff $\tilde{p}_i \le \alpha$ yields $\mathbb{P}(A_i = 1 \mid T_i = 0) \le \alpha$.
*Proof.* For $\alpha \in (0,1)$, $\{\tilde{p}_i \le \alpha\} = \{\rho_i \le \lfloor (N+1)\alpha \rfloor\}$, of null probability $\lfloor (N+1)\alpha \rfloor/(N+1) \le \alpha$ by Lemma 4.1.

This is the same rule as Theorem 4.1. The p-value formulation separates score computation from the decision rule. A verifier that rejects everything attains $\mathbb{P}(H_i) = 0$ trivially yet is useless, so the hallucination guarantee must be paired with a statement that true claims survive, which requires a complementary true-claim calibration set.

**Theorem 4.2 (Finite-sample false-rejection control for true claims).** Let $s_1^{\mathrm{true}}, \ldots, s_M^{\mathrm{true}}$ be calibration scores from known-true claims. Suppose that, conditional on $T_i = 1$, these scores and the test score $s_i$ are exchangeable, with randomized tie breaking if needed. Let

$$q_\beta = \lfloor (M+1)\beta \rfloor.$$

If $q_\beta = 0$, set $\eta_\beta = +\infty$; otherwise set

$$\eta_\beta = s_{(M+1-q_\beta)}^{\mathrm{true}}.$$

Under the rejection rule $s_i > \eta_\beta$,

$$P(s_i > \eta_\beta \mid T_i = 1) \le \frac{q_\beta}{M+1} \le \beta.$$

**Proof.** The randomized rank of $s_i$ among the $M+1$ true-class scores is uniform. If $q_\beta = 0$, rejection is impossible. Otherwise, rejection places the test score among the largest $q_\beta$ ranks, an event of probability at most $q_\beta/(M+1)$. ■

To combine this rule with the false-claim acceptance threshold $\tau_\alpha$, set

$$a = \min(\tau_\alpha, \eta_\beta), \qquad r = \max(\tau_\alpha, \eta_\beta),$$

accept when $s_i < a$, reject when $s_i > r$ and abstain otherwise. This conservative three-way rule preserves both one-sided bounds and converts any overlap of the unmodified rules into an abstention band.

**Proposition 4.3 (No nontrivial distribution-free false-acceptance control from true labels alone).** Let $T$ be a threshold measurable with respect to a true-claim calibration sample, and suppose a false test score $S_0$ is independent of that sample. If $T$ is almost surely finite, then

$$\sup_{F_0} P_{F_0}(S_0 < T) = 1.$$

Hence no uniform false-acceptance level $\alpha < 1$ can be guaranteed from true calibration alone. The trivial rule $T = -\infty$, which always abstains, is excluded.

**Proof.** Because $T$ is almost surely finite, $P(T > c) \to 1$ as $c \to -\infty$. For any $\varepsilon > 0$, choose a deterministic $c_\varepsilon$ with $P(T > c_\varepsilon) > 1 - \varepsilon$ and take $F_0$ to be a point mass at $c_\varepsilon$. Then

$$P_{F_0}(S_0 < T) = P(c_\varepsilon < T) > 1 - \varepsilon.$$

Let $\varepsilon \downarrow 0$. ■

Distribution-free hallucination control from a true-only calibration set is thus provably unattainable; one must supply either false-class labels or a separation assumption. The latter route is available when only true labels exist, at the cost of a non-distribution-free hypothesis.

**Theorem 4.3 (Detectability route: true calibration plus an explicit separation condition).** Use $\eta_\beta$ from Theorem 4.2. Assume that, conditional on $T_i = 0$, the false test score is independent of the true-calibration sample, with CDF $F_0$, and

$$F_0(\eta_\beta) \le \kappa \quad \text{almost surely.}$$

Then

$$P(s_i \le \eta_\beta \mid T_i = 0) = E\{F_0(\eta_\beta)\} \le \kappa.$$

If $F_0$ is estimated from an additional independent sample of $N$ false claims, then, with probability at least $1-\gamma$,

$$P(s_i \le \eta_\beta \mid T_i = 0) \le \hat{F}_0(\eta_\beta) + \sqrt{\frac{\log(2/\gamma)}{2N}}.$$

**Proof.** Conditional independence gives $P(s_i \le \eta_\beta \mid \eta_\beta, T_i = 0) = F_0(\eta_\beta)$. Taking expectations and applying the separation condition gives the first claim. The second follows from the uniform Dvoretzky–Kiefer–Wolfowitz inequality for the independent false-class sample.[4] ■

Assumption A5 is not distribution-free, it is exactly the detectability content Proposition 4.3 shows is indispensable when false labels are absent, so Theorem 4.1 remains the primary, distribution-free guarantee and Theorem 4.3 is the fallback whose price is the explicit separation assumption A5. The per-claim results lift to the whole report. Let $\mathcal{A} := \{i : A_i = 1\}$ be the accepted set and $N_H := \sum_{i=1}^{m} \mathbf{1}_{H_i} = \#\{i \in \mathcal{A} : T_i = 0\}$ the number of accepted hallucinations. By linearity of expectation, with no independence needed, under A1–A4 per claim $\mathbb{E}[N_H] = \sum_i \mathbb{P}(H_i) \le \alpha \sum_i \mathbb{P}(T_i = 0) \le \alpha\, m\, \pi_0$ with $\pi_0 := \max_i \mathbb{P}(T_i = 0)$; and by the union bound $\mathbb{P}(\cup_{i=1}^{m} H_i) \le \sum_i \mathbb{P}(H_i) \le \alpha\, m\, \pi_0$, so running each claim at level $\alpha = \alpha_{\text{rep}}/(m\pi_0)$ (or $\alpha_{\text{rep}}/m$ if $\pi_0$ is unknown) makes the report-wise false-acceptance probability at most $\alpha_{\text{rep}}$, a Bonferroni calibration. The sharper aggregate object is false-discovery control.

**Theorem 4.4 (Conditional FDR control for accepted false claims).** Let $\mathbf{T} = (T_1, \ldots, T_m)$ and $\mathcal{H}_0(\mathbf{T}) = \{i : T_i = 0\}$. Suppose that, conditional on every realized $\mathbf{T}$,

$$P(p_i \le u \mid \mathbf{T}) \le u, \qquad i \in \mathcal{H}_0(\mathbf{T}), \quad u \in [0,1].$$

If, conditional on $\mathbf{T}$, the full $p$-value vector is mutually independent, or is PRDS on the coordinates in $\mathcal{H}_0(\mathbf{T})$, the Benjamini–Hochberg procedure at level $q$ satisfies

$$\mathrm{FDR} \le E\left[\frac{m_0(\mathbf{T})}{m}\right] q \le q.$$

Under arbitrary conditional dependence, the Benjamini–Yekutieli procedure[5] at level $q/H_m$, where $H_m = \sum_{j=1}^{m} 1/j$, satisfies $\mathrm{FDR} \le q$.

**Proof.** Conditional on $\mathbf{T}$, the standard BH theorem applies under mutual independence or PRDS on the true-null subset, and the standard BY theorem applies under arbitrary dependence.[6] The null set is then fixed and its $p$-values are super-uniform by assumption. Taking expectation over $\mathbf{T}$ gives the unconditional bound. ■

Because claims in one report share evidence and an upstream generator, they are not independent by default. The BY form in part (b) provides the guaranteed bound under arbitrary dependence, whereas the BH form in part (a) describes the bound under a working independence or PRDS condition. The per-claim and FDR guarantees answer different questions: how often an accepted claim hallucinates, versus what fraction of accepted claims are hallucinated. Both guarantees are relevant for a multi-claim synthesis report, with the per-claim bound providing exact distribution-free control and the FDR bound providing the appropriate aggregate control.

The identity

$$P(H_i) = P(A_i = 1 \mid T_i = 0)P(T_i = 0) \le \alpha P(T_i = 0)$$

separates upstream generation from downstream verification. Note 4 controls only the conditional acceptance factor. Notes 2 and 3 analyze estimator bias and risk and do not establish that the implemented generator reduces $P(T_i = 0)$. Any reduction in the upstream false-claim base rate requires separate empirical or probabilistic analysis.

**Supplementary Note 5. Finite termination and residual upper bounds for iterative refinement**

This Note separates two questions: whether an abstract refinement process terminates under finite-progress conditions and what upper bounds follow if a non-negative residual satisfies a stochastic drift condition. The first is a well-founded progress result; the second controls an expected upper envelope and does not generally imply convergence when the additive drift allowance is positive. Within this Note, $\rho \in [0,1)$ denotes the geometric drift or contraction factor rather than the pairwise correlation in Note 2, and $b \geq 0$ is an additive drift allowance rather than a source bias.

For a fixed query, consider an abstract refinement process in rounds $t = 0,1, \ldots$. Its state is

$$x_t = (z_t, E_t, H_t, \Pi_t, U_t) \in \mathcal{X},$$

where $z_t$ is the query decomposition, $E_t$ the retrieved evidence, $H_t$ the claim set, $\Pi_t$ the provenance graph and $U_t$ the uncertainty/contradiction vector. The update may be stochastic and time-varying,

$$x_{t+1} = \Psi_t(x_t, \omega_{t+1}), \qquad \mathcal{F}_t = \sigma(x_0, \omega_1, \ldots, \omega_t).$$

The results below treat finite progress, stochastic drift and deterministic contraction as explicit sufficient conditions rather than as unconditional implementation facts.

$$V(x) := w_U \parallel U \parallel_1 + w_K \,\mathrm{Contra}(H, \Pi) + w_G \,\mathrm{Gap}(H, \Pi) \ \geq 0,$$

Here $\parallel U \parallel_1$ is total residual uncertainty, $\mathrm{Contra}(H, \Pi)$ is the contradiction mass among accepted claims flagged by the screening procedure and $\mathrm{Gap}(H, \Pi)$ is the ungrounded-claim mass. Thus $V(x) \geq 0$ is a code- or model-defined residual, not a measure of factual or clinical correctness. The quantity $b \geq 0$ below is an additive drift allowance; it need not be the smallest admissible constant and is not, by itself, a lower bound or limiting residual.

The results use three separate assumption layers.

**A1 (Finite admissible refinement space).** For a fixed query, there is a finite set $\mathcal{A}$ of admissible refinement opportunities, with $|\mathcal{A}| = L < \infty$.

**A2 (No repetition).** Let $Q_t \subseteq \mathcal{A}$ be the opportunities completed by the end of round $t$. Then $Q_t \subseteq Q_{t+1}$, and a completed opportunity is not used again.

**A3 (Progress or stop).** The process stops if $V_t \leq \epsilon$ or $Q_t = \mathcal{A}$. Otherwise, the next round consumes at least one previously unused opportunity:

$$|Q_{t+1}| \geq |Q_t| + 1. \tag{P}$$

This condition does not assert that $V_t$ strictly decreases.

**A4 (Stochastic drift).** There exist $0 \leq \rho < 1$ and $b \geq 0$ such that

$$E[V_{t+1} \mid \mathcal{F}_t] \leq \rho V_t + b \quad \text{almost surely.} \tag{5.1}$$

**A5 (Deterministic contraction).** The update is deterministic and time-homogeneous on a complete metric state space and is a contraction with factor $\rho < 1$.

The finite-termination theorem uses A1–A3, the residual upper bound uses A4 and the Banach result uses A5. None of these layers is inferred from another.

The first primary result is that the loop provably halts.

**Theorem 5.1 (Finite termination under finite-progress conditions).** Under A1–A3, the refinement process terminates after at most

$$T \leq |\mathcal{A} \backslash Q_0|$$

update rounds. At termination, either $V_T \le \epsilon$ or $Q_T = \mathcal{A}$, in which case every admissible refinement opportunity has been used. This is a conditional sufficient-condition theorem; it does not assert that a particular implementation satisfies A1–A3. It also does not imply $V_T = 0$, correctness of the terminal report or uniqueness of the terminal state.

**Proof.** Define the integer-valued potential

$$\Phi_t = |\mathcal{A} \backslash Q_t| \in \{0,1,\dots,L\}.$$

By A2, $\Phi_{t+1} \le \Phi_t$. By A3, every non-terminal round satisfies

$$\Phi_{t+1} \le \Phi_t - 1.$$

A non-negative integer can decrease strictly at most $\Phi_0 = |\mathcal{A} \backslash Q_0|$ times. The stopping rule therefore fires within the displayed number of updates, at either the residual or refinement-exhaustion condition. ■

**Theorem 5.2 (Geometric upper bound for the expected synthesis residual).** Assume A4, let $V_t = V(x_t) \ge 0$ and suppose $E[V_0] < \infty$. Then:
(a) for every $t \ge 0$,

$$E[V_t] \le \rho^t E[V_0] + b\,\tfrac{1-\rho^t}{1-\rho} \le \rho^t E[V_0] + \tfrac{b}{1-\rho}; \tag{5.2}$$

(b)

$$\limsup_{t\to\infty} E[V_t] \le \frac{b}{1-\rho};$$

(c) for every $\delta \in (0,1)$,

$$P\left[V_t > \frac{1}{\delta}\left\{\rho^t E[V_0] + b\,\frac{1-\rho^t}{1-\rho}\right\}\right] \le \delta;$$

(d) if $b = 0$, then

$$E[V_t] \le \rho^t E[V_0] \to 0, \qquad \sum_{t=0}^{\infty} V_t < \infty \text{ a.s.}, \qquad V_t \to 0 \text{ a.s.}$$

**Proof.** Taking total expectations in A4 gives $E[V_{t+1}] \le \rho E[V_t] + b$. Induction yields

$$E[V_t] \le \rho^t E[V_0] + b\sum_{j=0}^{t-1} \rho^j = \rho^t E[V_0] + b\,\frac{1-\rho^t}{1-\rho},$$

which proves part (a). Letting $t \to \infty$ proves part (b), and part (c) follows from Markov's inequality. If $b = 0$, Tonelli's theorem gives

$$E\left[\sum_{t=0}^{\infty} V_t\right] = \sum_{t=0}^{\infty} E\,[V_t] \le \frac{E[V_0]}{1-\rho} < \infty.$$

Hence the non-negative series is finite almost surely, and therefore $V_t \to 0$ almost surely. ■

Let $B = b/(1-\rho)$. If $0 < \rho < 1$ and $E[V_0] > \epsilon > B$, then the simpler envelope in (5.2) is at most $\epsilon$ whenever

$$t \geq \left\lceil \frac{\log\{E[V_0]/(\epsilon - B)\}}{\log(1/\rho)} \right\rceil.$$

If $E[V_0] \leq \epsilon$, zero additional rounds are needed; if $\rho = 0$, one round suffices for every $\epsilon > b$. For $\epsilon \leq B$, the upper bound is non-certifying: it does not imply that the actual residual cannot fall below $\epsilon$. When $b > 0$, A4 alone implies neither convergence of $E[V_t]$ nor convergence, monotonicity or a positive lower bound for $V_t$. Only the transient term in the upper envelope decays geometrically.

**Lemma 5.1 (A stagewise sufficient condition for zero-additive-drift contraction).** Write one deterministic post-saturation round as

$$\Psi = F_{\text{refine}} \circ F_{\text{verify}} \circ F_{\text{upstream}}.$$

Suppose, as separate assumptions about the same residual $V$, that

$$V(F_{\text{upstream}}x) \leq V(x), \qquad V(F_{\text{verify}}x) \leq V(x), \qquad V(F_{\text{refine}}x) \leq \rho V(x)$$

for every admissible input and some $0 \leq \rho < 1$. Then

$$V(\Psi x) \leq \rho V(x).$$

**Proof.**

$$V(\Psi x) \leq \rho V(F_{\text{verify}}(F_{\text{upstream}}x)) \leq \rho V(F_{\text{upstream}}x) \leq \rho V(x).$$

■

This is an independent sufficient-condition decomposition. Note 4 controls a false-acceptance probability and does not establish non-expansiveness of the distinct Note-5 residual.

**Theorem 5.3 (Banach fixed point—deterministic special case).** Under A5: (i) $\Psi$ is a contraction; (ii) there is a unique fixed point $x^\star = \Psi x^\star$; (iii) $x_t = \Psi^t x_0 \to x^\star$ from any $x_0$; (iv) the rate is geometric,

$$d(x_t, x^\star) \leq \rho^t d(x_0, x^\star);$$

and (v) the a posteriori bounds

$$d(x_t, x^\star) \leq \frac{\rho^t}{1-\rho} d(x_1, x_0), \qquad d(x_t, x^\star) \leq \frac{1}{1-\rho} d(x_t, x_{t+1})$$

hold.

**Proof.** The contraction premise is A5. Completeness and the Banach fixed-point theorem give existence, uniqueness and convergence. Iterating the contraction inequality gives

$$d(x_{k+1}, x_k) \leq \rho^k d(x_1, x_0).$$

For $n > t$,

$$d(x_t, x_n) \le \sum_{k=t}^{n-1} \rho^k \, d(x_1, x_0) \le \frac{\rho^t}{1-\rho} d(x_1, x_0),$$

so $\{x_t\}$ is Cauchy and converges to some $x^\star$. Because $\Psi$ is Lipschitz, it is continuous and $x^\star = \Psi x^\star$. Uniqueness follows from

$$d(y, y') = d(\Psi y, \Psi y') \le \rho d(y, y')$$

for two fixed points and $\rho < 1$. The first rate follows by induction; the a posteriori bounds follow by taking $n \to \infty$ in the Cauchy estimate and by rearranging

$$d(x_t, x^\star) \le d(x_t, x_{t+1}) + \rho d(x_t, x^\star).$$

■

Theorem 5.3 is a separate deterministic result under A5, not a limiting consequence of the stochastic drift condition A4. The weighted metric is used only for this deterministic result; the finite-progress and drift results require no metric on $\mathcal{X}$. In the deterministic contraction regime, $x_t \to x^\star$ in $d$. If $V$ is continuous with respect to $d$, then $V(x_t) \to V(x^\star)$; continuity is an additional requirement for this scalar conclusion.

The drift result is robust to bounded time inhomogeneity. If

$$E[V_{t+1} \mid \mathcal{F}_t] \le \rho_t V_t + b_t, \qquad \sup_t \rho_t \le \rho < 1, \qquad \sup_t b_t \le b,$$

then replacing $\rho_t, b_t$ by their suprema gives the same upper envelope because $V_t \ge 0$. A post-saturation analysis may begin at any stopping time after which the evidence component is fixed, but finiteness of an evidence universe alone does not imply saturation by a prescribed round count.

Note 5 controls termination under A1–A3, an expected residual upper envelope under A4 and a deterministic fixed point under A5. The residual $V$ is not a correctness measure: a low-residual state may still reflect incomplete, biased or inapplicable evidence. Notes 2–3, Note 4 and Note 5 concern different mathematical objects. Notes 2–3 compare idealized estimators at a fixed query; Note 4 controls a calibrated false-acceptance probability; Note 5 analyzes an iterative residual. No theorem identifies the terminal workflow state with the estimator optimized in Notes 2–3, and neither Note 4 nor Notes 2–3 supplies a Note-5 contraction assumption.

**Supplementary Note 6. Finite-sample generalization of multi-source synthesis**

This Note asks whether, when a synthesis function is learned from finitely many labeled examples drawn from heterogeneous sources, its risk on the deployment target distribution can be controlled, and what that implies for how the sources should be weighted. Within this Note $R$ (and $R_P$, $R_w$, $R_\star$, $\hat{R}_w$) denotes the population or empirical risk of a synthesis function $\mathbb{E}[\ell(f(X), Y)]$ — not the MSE estimation risk of Notes 2 and 3; the two are the same notion of expected loss but act on different objects, an aggregated estimator there and a learned function here. Here $\delta \in (0,1)$ is the failure probability of the high-probability bound, by the learning-theory convention, appearing only as $\log(1/\delta)$ and unsubscripted, hence distinct from the idiosyncratic bias $\delta_s$ of Notes 2 and 3; $\lambda_w$ is the joint optimal risk, consistent with Note 3; and $w \in \Delta^{K-1}$ are source weights as before.

Let $\mathcal{X}$ be the input space and $\mathcal{Y}$ the label space. Sources $s \in \{1, \ldots, K\}$ induce distributions $P_s$ on $\mathcal{X} \times \mathcal{Y}$, the target distribution is $P_\star$, the hypothesis class is $\mathcal{F}$ and the bounded loss is $\ell: \mathcal{Y} \times \mathcal{Y} \rightarrow [0, M]$. Write

$$R_P(f) = E_{(X,Y)\sim P}\ell\{f(X), Y\}, \qquad P_w = \sum_s w_s P_s, \qquad R_w(f) = \sum_s w_s R_s(f).$$

From source $s$, observe $S_s = \{Z_{s,i}\}_{i=1}^{n_s}$ iid from $P_s$, with source samples independent across $s$, and define

$$\hat{R}_w(f) = \sum_s \frac{w_s}{n_s} \sum_{i=1}^{n_s} \ell\{f(X_{s,i}), Y_{s,i}\}.$$

Then $E[\hat{R}_w(f)] = R_w(f)$.

**Applicability boundary.** The finite-sample results apply when the objects indexed by source are independent labeled training examples from the stated distributions. Evidence documents, estimated study effects, EHR cohorts and knowledge-graph relations are not automatically iid observations satisfying A4. A concrete MES deployment is covered only after specifying the learned function, label, sampling unit and independence structure that instantiate this model.
For the loss class

$$\mathcal{G} = \{(x, y) \mapsto \ell(f(x), y) : f \in \mathcal{F}\},$$

define the empirical weighted Rademacher complexity

$$\hat{\mathfrak{R}}_w(\mathcal{G}; S) := E_\sigma\left[\sup_{g\in\mathcal{G}} \sum_{s=1}^{K} \frac{w_s}{n_s} \sum_{i=1}^{n_s} \sigma_{s,i}\, g(Z_{s,i}) \,\middle|\, S\right], \tag{6.1}$$

where the $\sigma_{s,i}$ are independent Rademacher signs. Its expectation[7] over source samples is

$$\mathfrak{R}_w(\mathcal{G}) := E_S[\hat{\mathfrak{R}}_w(\mathcal{G}; S)]. \tag{6.2}$$

The hat always denotes the observed, sample-dependent quantity. Also define

$$n_{\text{eff}}(w) = \left(\sum_{s=1}^{K} \frac{w_s^2}{n_s}\right)^{-1}.$$

The label-aware loss-class discrepancy[8] is

$$\mathrm{disc}_{\mathcal{G}}(P,Q) = \sup_{g\in\mathcal{G}} |E_P g - E_Q g|, \qquad D_w(P_\star) = \sum_s w_s \,\mathrm{disc}_{\mathcal{G}}(P_s, P_\star).$$

The standing assumptions are bounded loss (A1: $0 \le \ell \le M < \infty$, satisfied by 0–1 loss, clipped log-loss, Brier score), measurability and mild regularity (A2: $(x,y) \mapsto \ell(f(x),y)$ measurable and $\mathcal{F}$ pointwise-measurable so that suprema over $\mathcal{G}$ are measurable and Fubini applies in symmetrization), convex source weights (A3: $w \in \Delta^{K-1}$), and independent source samples (A4: $S_1, \ldots, S_K$ mutually independent, within source i.i.d. from $P_s$). Not assumed are realizability ($\inf_f R_\star(f)$ may exceed 0), a covariate-shift-only restriction for the loss-IPM bound, equal sample sizes, or input–label independence; a joint term enters only in the label-agnostic route, flagged there. The argument proceeds in three layers, i.e., population transfer, finite-sample deviation, and the ERM oracle inequality. The first layer rests on the convexity of the discrepancy in the weights: since $\sum_s w_s = 1$, for any $g \in \mathcal{G}$ one has $\mathbb{E}_{P_w} g - \mathbb{E}_{P_\star} g = \sum_s w_s \left(\mathbb{E}_{P_s} g - \mathbb{E}_{P_\star} g\right)$, and taking absolute values and a supremum over $g$ gives the sub-additivity $\mathrm{disc}_{\mathcal{G}}(P_w, P_\star) \le \sum_s w_s D_s = D_w(P_\star)$. The population transfer inequality follows at once in its tight, label-aware form.

**Theorem 6.A (Loss-IPM transfer — tight, no joint term).** Under A1–A3, for every fixed $f$ and $w \in \Delta^{K-1}$, $|R_\star(f) - R_w(f)| \le \mathrm{disc}_{\mathcal{G}}(P_w, P_\star) \le D_w(P_\star)$, hence $R_\star(f) \le R_w(f) + D_w(P_\star)$, and no $\lambda_w$ term is needed or helpful.

*Proof.* $R_\star(f) - R_w(f) = \mathbb{E}_{P_\star} g_f - \mathbb{E}_{P_w} g_f$ with $g_f \in \mathcal{G}$; bound by the integral-probability-metric definition, then by the sub-additivity just established.

**Binary label-agnostic special case.** This route is stated only for binary classification, $\mathcal{H} \subseteq \{0,1\}^{\mathcal{X}}$, and 0–1 risk. Define

$$d_{\mathcal{H}\Delta\mathcal{H}}(P_X, Q_X) = 2 \sup_{h,h'\in\mathcal{H}} |P_X\{h(X) \ne h'(X)\} - Q_X\{h(X) \ne h'(X)\}|$$

and

$$\lambda_w = \inf_{h\in\mathcal{H}} \{R_w(h) + R_\star(h)\}.$$

Then the standard binary domain-adaptation inequality[9] gives

$$R_\star(h) \le R_w(h) + \frac{1}{2} d_{\mathcal{H}\Delta\mathcal{H}}(P_{w,X}, P_{\star,X}) + \lambda_w,$$

with

$$d_{\mathcal{H}\Delta\mathcal{H}}(P_{w,X}, P_{\star,X}) \le \sum_s w_s \, d_{\mathcal{H}\Delta\mathcal{H}}(P_{s,X}, P_{\star,X}).$$

This statement does not apply as written to Brier or clipped log loss; Theorem 6.A supplies the label-aware loss-IPM route for those losses.

**Proof.** Apply the binary $\mathcal{H}\Delta\mathcal{H}$ domain-adaptation inequality to $P_w$, then use linearity under the mixture, the triangle inequality and the supremum to obtain the second display. ∎

For the finite-sample layer, changing one source-$s$ observation changes a fixed weighted empirical risk by at most $w_s M / n_s$. McDiarmid's inequality[10] therefore has squared bounded-difference sum

$$M^2 \sum_s \frac{w_s^2}{n_s} = \frac{M^2}{n_{\mathrm{eff}}(w)}.$$

The effective-sample-size term alone cannot be smaller than the pooled equal-observation contribution because $n_{\mathrm{eff}}(w) \le \sum_s n_s$ . Weighting nevertheless also changes empirical risk, discrepancy and empirical complexity; the full certificate, not the variance term alone, must be compared.

**Lemma 6.1 (Expected- and empirical-complexity deviation bounds).** Under A1–A4, for any fixed $w \in \Delta^{K-1}$:

(a) with probability at least $1-\delta$,

$$\sup_{f\in\mathcal{F}}\{R_w(f) - \hat{R}_w(f)\} \le 2\mathfrak{R}_w(\mathcal{G}) + M\sqrt{\frac{\log(1/\delta)}{2n_{\mathrm{eff}}(w)}}; \tag{6.3}$$

(b) with probability at least $1-\delta$,

$$\sup_{f\in\mathcal{F}}\{R_w(f) - \hat{R}_w(f)\} \le 2\hat{\mathfrak{R}}_w(\mathcal{G}; S) + 3M\sqrt{\frac{\log(2/\delta)}{2n_{\mathrm{eff}}(w)}}; \tag{6.4}$$

(c) with probability at least $1-\delta$,

$$\sup_{f\in\mathcal{F}}|R_w(f) - \hat{R}_w(f)| \le 2\hat{\mathfrak{R}}_w(\mathcal{G}; S) + 3M\sqrt{\frac{\log(4/\delta)}{2n_{\mathrm{eff}}(w)}}. \tag{6.5}$$

**Proof.** Set

$$\Phi(S) = \sup_{g\in\mathcal{G}}\left\{E_{P_w}g - \sum_s \frac{w_s}{n_s}\sum_i g\,(Z_{s,i})\right\}.$$

Replacing one source-$s$ observation changes $\Phi$ by at most $w_s M/n_s$, so the sum of squared bounded differences is $M^2/n_{\mathrm{eff}}(w)$. McDiarmid's inequality gives, with probability at least $1-\delta$,

$$\Phi(S) \le E\Phi(S) + M\sqrt{\frac{\log(1/\delta)}{2n_{\mathrm{eff}}(w)}}.$$

Ghost-sample symmetrization yields $E\Phi(S) \le 2\mathfrak{R}_w(\mathcal{G})$, proving part (a).

The map $S \mapsto \hat{\mathfrak{R}}_w(\mathcal{G}; S)$ has the same bounded differences. Thus, with probability at least $1-\delta/2$,

$$\mathfrak{R}_w(\mathcal{G}) \le \hat{\mathfrak{R}}_w(\mathcal{G}; S) + M\sqrt{\frac{\log(2/\delta)}{2n_{\mathrm{eff}}(w)}}.$$

Combining this event with the first concentration event at failure probability $\delta/2$ gives the coefficient $3M$ in (6.4). For the reverse deviation, apply (6.4) to $M-\mathcal{G}$; its empirical Rademacher complexity equals that of $\mathcal{G}$ by sign symmetry. A union bound over the two tails gives (6.5). ■

Chaining the population and finite-sample layers yields the headline generalization bound.

**Theorem 6.B (Fixed-weight finite-sample target-risk bound).** Under A1–A4, let $w \in \Delta^{K-1}$ be fixed independently of the observed source samples. With probability at least $1-\delta$, simultaneously for every $f \in \mathcal{F}$,

$$R_\star(f) \le \hat{R}_w(f) + D_w(P_\star) + 2\widehat{\mathfrak{R}}_w(\mathcal{G}; S) + 3M\sqrt{\frac{\log(2/\delta)}{2n_{\mathrm{eff}}(w)}}. \tag{6.6}$$

The theorem remains valid if the population discrepancy is replaced by a simultaneous upper confidence bound. It is not automatically fully observable because $D_w(P_\star)$ may require labeled target data.

**Proof.** Theorem 6.A gives $R_\star(f) \le R_w(f) + D_w(P_\star)$. Lemma 6.1(b) bounds $R_w(f)$ by the remaining right-hand-side terms uniformly over $f$. Chaining the inequalities proves (6.6). ■

**Theorem 6.C (Oracle inequality for approximate weighted ERM).** Fix $w$ independently of the observed samples. Let $\hat{f}_w$ be an $\varepsilon_{\mathrm{opt}}$-empirical minimizer:

$$\hat{R}_w(\hat{f}_w) \le \inf_{f\in\mathcal{F}} \hat{R}_w(f) + \varepsilon_{\mathrm{opt}},$$

and define

$$\Gamma_w(\delta) = 2\widehat{\mathfrak{R}}_w(\mathcal{G}; S) + 3M\sqrt{\frac{\log(4/\delta)}{2n_{\mathrm{eff}}(w)}}.$$

Then, with probability at least $1-\delta$,

$$R_\star(\hat{f}_w) \le \inf_{f\in\mathcal{F}} R_\star(f) + 2\Gamma_w(\delta) + 2D_w(P_\star) + \varepsilon_{\mathrm{opt}}. \tag{6.7}$$

**Proof.** On the event $\sup_f |R_w(f) - \hat{R}_w(f)| \le \Gamma_w$, choose $f_\eta$ with $R_\star(f_\eta) \le \inf_f R_\star(f) + \eta$. Then

$$R_\star(\hat{f}_w) \le R_w(\hat{f}_w) + D_w.$$

Next,

$$R_w(\hat{f}_w) + D_w \le \hat{R}_w(\hat{f}_w) + \Gamma_w + D_w \le \hat{R}_w(f_\eta) + \varepsilon_{\mathrm{opt}} + \Gamma_w + D_w.$$

Finally,

$$\hat{R}_w(f_\eta) + \varepsilon_{\mathrm{opt}} + \Gamma_w + D_w \le R_w(f_\eta) + \varepsilon_{\mathrm{opt}} + 2\Gamma_w + D_w \le R_\star(f_\eta) + \varepsilon_{\mathrm{opt}} + 2\Gamma_w + 2D_w.$$

Letting $\eta \downarrow 0$ proves (6.7). ■

The oracle inequality contains estimation, optimization and source–target-discrepancy penalties. The term $2D_w(P_\star)$ is an upper-bound penalty, not a proved lower bound or irreducible "shift floor" for the actual excess risk.

**Corollary 6.C.1 (Valid post-selection from a finite weight library).** Let

$$\mathcal{W}_0 = \{w^{(1)}, \ldots, w^{(L)}\} \subset \Delta^{K-1}$$

be fixed independently of the observed samples. With probability at least $1-\delta$, simultaneously for every $\ell = 1, \ldots, L$ and $f \in \mathcal{F}$,

$$\mathcal{C}_\ell(f) := \hat{R}_{w^{(\ell)}}(f) + D_{w^{(\ell)}}(P_\star) + 2\widehat{\mathfrak{R}}_{w^{(\ell)}}(\mathcal{G}; S).$$

On the same event,

$$R_\star(f) \le \mathcal{C}_\ell(f) + 3M\sqrt{\frac{\log(2L/\delta)}{2n_{\mathrm{eff}}(w^{(\ell)})}}. \tag{6.8}$$

Thus, a candidate may be selected after observing the sample by minimizing the full right-hand side over the fixed library. The selected point may be a vertex, boundary point or interior mixture. Continuous adaptive optimization requires an independent tuning sample or a separate uniform-in-$w$ theorem.

**Proof.** Apply Theorem 6.B to each candidate at failure probability $\delta/L$ and take a union bound. ■

Reliability reweighting is not automatically beneficial. For $w \in \mathcal{W}_0$, define

$$C_\delta(w) = \hat{R}_w(\hat{f}_w) + D_w(P_\star) + 2\widehat{\mathfrak{R}}_w(\mathcal{G}; S) + 3M\sqrt{\frac{\log(2L/\delta)}{2n_{\mathrm{eff}}(w)}}.$$

Reweighting to $\widetilde{w}$ tightens the certificate exactly when

$$C_\delta(\widetilde{w}) \le C_\delta(w);$$

every empirical-risk, discrepancy, complexity and effective-sample-size term must be included. If a screening or weighting rule generates weights from the same data outside a prespecified finite library, the fixed-weight theorem does not cover the selected weight without sample splitting or a uniform-in-$w$ result.

### Supplementary Note 7. Direct-prompting baseline for CliniFact verification

For the direct-prompting baseline used in the CliniFact evaluation, we applied the original three-class classification prompt to each claim-evidence pair. The model was given one scientific claim and one abstract and was instructed to classify whether the abstract supported the claim, reported inconclusive findings, or provided no relevant information. The output was constrained to a JSON object with RESULT as the key. The prompt template was as follows, with {claim} and {abstract} replaced by the corresponding claim and source abstract for each evaluation instance.

```
Given a scientific claim and an abstract, determine if the abstract reports positive results
(TRUE), inconclusive results (FALSE), or offers no information (NONE) about the claim.

The task is to classify the pair claim abstract as follows:
TRUE: if the abstract provide support for the claim.
FALSE: if the abstract provide inconclusive support for the claim.
NONE: if the abstract provides contextual or background information without directly reporting
results about the claim.
Claim: {claim}
Abstract: {abstract}

Output a json dict with RESULT as the key. output a single JSON string inline (not in a code
block).
```

For analysis, TRUE predictions were mapped to the Evidence class, FALSE predictions were mapped to the Inconclusive class and NONE predictions were mapped to the not enough information (NEI) class. In the binary unsupported-claim detection analysis, FALSE and NONE were grouped as non-supporting evidence.
